# Combatting seasonal malaria transmission using a highly potent *Plasmodium falciparum* transmission-blocking monoclonal antibody

**DOI:** 10.1101/2022.09.11.22279612

**Authors:** Joseph D Challenger, Stijn W van Beek, Rob ter Heine, Saskia C van der Boor, Giovanni D Charles, Merel J Smit, Chris Ockenhouse, John J Aponte, Matthew BB McCall, Matthijs M Jore, Thomas S Churcher, Teun Bousema

## Abstract

Transmission-blocking interventions can play an important role in combatting malaria worldwide. Recently, a highly potent *Plasmodium falciparum* transmission-blocking monoclonal antibody (TB31F) was demonstrated to be safe and efficacious in malaria-naïve volunteers. Here we determine what dose would be required to obtain effective transmission reduction throughout the malaria season and predict the potential public health impact of large-scale implementation of TB31F alongside existing interventions. To this purpose, we developed a pharmaco-epidemiological model, tailored to two settings of differing transmission intensity with already established insecticide-treated nets and seasonal malaria chemoprevention interventions. We found that a simple weight-based TB31F dosing strategy achieved >80% transmission-reducing activity for over 5 months. With this approach, community-wide annual administration (at 80% coverage) of TB31F over a three-year period was predicted to reduce clinical incidence by 54% (381 cases averted per 1000 people per year) in a high-transmission seasonal setting, and 74% (157 cases averted per 1000 people per year) in a low-transmission seasonal setting. Targeting school-aged children gave the largest reduction in terms of cases averted per dose. We conclude that annual administration of transmission-blocking mAb TB31F may be an effective intervention against malaria in seasonal malaria settings.

**Key Questions:** *What is already known on this topic:*

- Naturally acquired transmission reducing antibodies can prevent malaria transmission to mosquitoes
- The humanized transmission-blocking monoclonal antibody TB31F is safe and single dose administration can achieve antibody concentrations that prevent malaria transmission for at least 84 days

*What this study adds:*

- A simple dosing regimen of TB31F in three weight-bands allows for single dose administration to sustain >80% transmission reducing activity for >5 months
- Community administration of TB31F can prevent a similar number of clinical malaria episodes compared to the highly efficacious seasonal malaria chemoprevention
- Community impact can be maximized when TB31F is combined with seasonal malaria chemoprevention
- School age children are the most effective part of the population to be targeted for maximum impact

*How this study might affect research, practice or policy:*

- Transmission blocking monoclonal antibodies can have a profound effect on malaria burden and can be combined with current interventions for maximum impact
- The predicted community impact of TB31F supports further clinical development of transmission-blocking monoclonal antibodies and exploration of use scenarios

## Introduction

Malaria remains a major health problem and caused approximately 627,000 deaths in 2020 (1). Considerable progress has been made in reducing the burden of malaria (2) and the recent recommendation of the World Health Organization to implement vaccination with the first-ever malaria vaccine RTS,S/AS01 fuels optimism for further successes. At the same time, progress towards malaria eradication has slowed in recent years (1) and the emergence and spread of insecticide and drug resistance threaten the efficacy of the interventions that were responsible for much of the recent progress in malaria control (3,4). New interventions that reduce the transmission of malaria are high on the priority list of tools for malaria control and eradication (5), and can play an important role in the containment of drug-resistant malaria.

Transmission-blocking vaccines aim to elicit antibodies that interfere with the transmission of malaria to mosquitoes by preventing fertilization of *Plasmodium* transmission stages, gametes, or later sporogonic development in the mosquito gut (6). The immediate consequence of vaccination is an antibody-mediated reduction in the infection (oocyst) burden in mosquitoes. The reduction in oocyst density is known as transmission-reducing activity (TRA). TRA is closely associated with the reduction in the proportion of mosquitoes that become infected (transmission-blocking activity, TBA) (7), although this relationship varies with oocyst intensity in control mosquitoes. TRA is more commonly reported in mosquito feeding assay experiments as, unlike TBA, its quantification is consistent across different experiments. However, TBA is the more relevant metric to describe efficacy in the field since the number of infected mosquitoes that is averted is more closely related to the ultimate public health outcome (8). By reducing the number of infected mosquitoes, transmission-blocking vaccines reduce malaria incidence at community level. Transmission-blocking vaccines based on pre-fertilization gametocyte antigens Pfs230 and Pfs48/45 are currently in clinical trials (6,9). These vaccines may be deployed as stand-alone interventions or in combination with anti-infection vaccines to increase their community impact (10). Instead of inducing antibodies by active immunisation, monoclonal antibodies (mAb) targeting the same antigens may be administered directly to achieve the same impact.

TB31F is a humanised version of the highly potent transmission-blocking rat mAb 85RF45.1 (11–14). It targets a highly conserved epitope on Pfs48/45, which is expressed on *Plasmodium falciparum* mature gametocytes and gametes. In a recent first-in-human study, a single intravenous dose up to 10 mg/kg was well-tolerated in adult study participants with minimal to no side-effects (15). In the highest dose group, serum from trial participants fully prevented transmission to mosquitoes in *ex-vivo* assays throughout 84 days of follow-up. Extrapolation based on the estimated antibody elimination half-life suggests that the effective activity following a single administration of TB31F could span a large proportion of the malaria transmission season in areas of seasonal malaria (16). This profile of activity makes the intervention attractive from an implementation perspective. It is currently unclear what dose would be required to obtain effective transmission reduction throughout the malaria season, what the expected community impact would be, and what sections of the population need to be targeted for maximum efficiency of the intervention. Demographically targeted interventions are attractive from an operational perspective but, unlike interventions that provide direct personal protection, require an understanding of what populations are most important for sustaining onward transmission to mosquitoes (17,18).

The aim of this study was to predict the potential public health impact of different implementation strategies with the transmission-blocking mAb TB31F. We combined a pharmacological model describing the TB31F exposure-response relationship with a modelling framework that captures the impact of changes in the human infectious reservoir for malaria on (clinical) malaria incidence. This pharmaco-epidemiological model allowed us to predict the impact of a large-scale intervention with TB31F alongside established public health interventions against malaria, in two highly seasonal malaria settings with markedly different transmission intensity.

## Materials and Methods

### Data

We used data from a first-in-human, dose escalation study of TB31F that was performed in 25 healthy adult malaria naïve volunteers (15). There were five study arms (n=5 per arm) with escalating TB31F dose: four groups received intravenous TB31F at 0.1 mg/kg, 1 mg/kg, 3 mg/kg and 10 mg/kg; a fifth group received 100 mg TB31F subcutaneously. Serum for pharmacokinetic and pharmacodynamic analysis was collected prior to administration, upon the end of infusion, at 1, 3, and 6 hours and on days 1, 2, 7, 14, 21, 28, 56, and 84 after the end of administration. TB31F serum concentrations were quantified by an ELISA against the recombinant protein R0.6C that contains the 6C fragment of the Pfs48/45 antigen to which TB31F binds. Transmission-reducing activity was determined by standard membrane feeding assay (SMFA) with cultured *P. falciparum* gametocytes and laboratory-reared *Anopheles stephensi* mosquitoes (19,20). The SMFA measured the reduction of oocyst density in mosquitoes fed on gametocytes in the presence of participants’ serum compared to pooled naïve serum. One participant in the subcutaneous group was excluded from all analyses due to implausibly low TB31F concentrations, potentially following incorrect administration.

### Pharmacokinetic/pharmacodynamic modelling

Parametric nonlinear mixed-effects modelling was performed using NONMEM version 7.4.1 to analyse the pharmacokinetic data and the relationship with transmission-reducing activity (21) as described in detail in the Supplemental Methods. We developed a practical weight-based intravenous dosing regimen reaching an equivalent duration >80% TRA in children as observed with the highest dose tested in adults using simulations with the pharmacokinetic/pharmacodynamic model, implementing an age-dependent exponent for allometric scaling of pharmacokinetic clearance parameters (Supplementary Methods) (22). Weight-based dosing regimens were explored aiming to reach a time to 80% transmission-reducing activity that was similar to that estimated from the trial data for a 70 kg adult administered 10 mg/kg TB31F intravenously.

### Predicting public health impact using transmission modelling

To gain insight into the potential epidemiological impact of this novel intervention, we incorporated the results of the pharmacokinetic-pharmacodynamic model into a mathematical model of malaria transmission. We have used a stochastic, individual-based implementation of the model originally outlined by Griffin et al. (23,24) which incorporates heterogeneity of transmission, naturally acquired immunity to infection, and is calibrated to reproduce age-dependent patterns of malaria incidence and prevalence from multiple sites across sub-Saharan Africa. The model also allows the introduction of interventions against malaria, such as ITNs, treatment for symptomatic disease, and SMC. This transmission model is described in detail elsewhere (23,24). Briefly, the malaria status of humans can transition between 6 states (Figure 1, panel 7). When uninfected individuals (*S*) are infected by a feeding Anopheles mosquito, the infection can either become symptomatic or remain asymptomatic (*A*), depending on their degree of naturally acquired immunity against malaria. Symptomatic infections are either treated with antimalarials (*T*) or remain untreated (*D*), which will depend on the level of treatment coverage selected in the model. If the infection is treated, the individual benefits from a period of prophylaxis (*P*), during which they are protected against further infectious bites. The individual then returns to being malaria-susceptible (*S*). If the symptomatic infection is untreated, the infection will become a long-lasting asymptomatic infection (*A*). Eventually, the parasite densities in these infections will be reduced due to the human immune response, and the infection becomes undetectable by slide microscopy (*U*). When the infection is cleared by the immune response, the individual returns to being malaria-susceptible (*S*). Humans in states *D, T, A*, and *U* are able to transmit malaria to feeding mosquitos, although not all infected humans are equally infectious, those with sub-microscopic infections having the lowest per-bite probability of infecting a mosquito. The mosquito component of the model is simpler: mosquitoes are either susceptible to malaria (*S*_*M*_), infected but not yet infectious (*E*_*M*_), or infectious (*I*_*M*_). The size of the adult female mosquito population, relative to that of the human population, determines the baseline intensity of transmission in the model.

**Figure 1.**
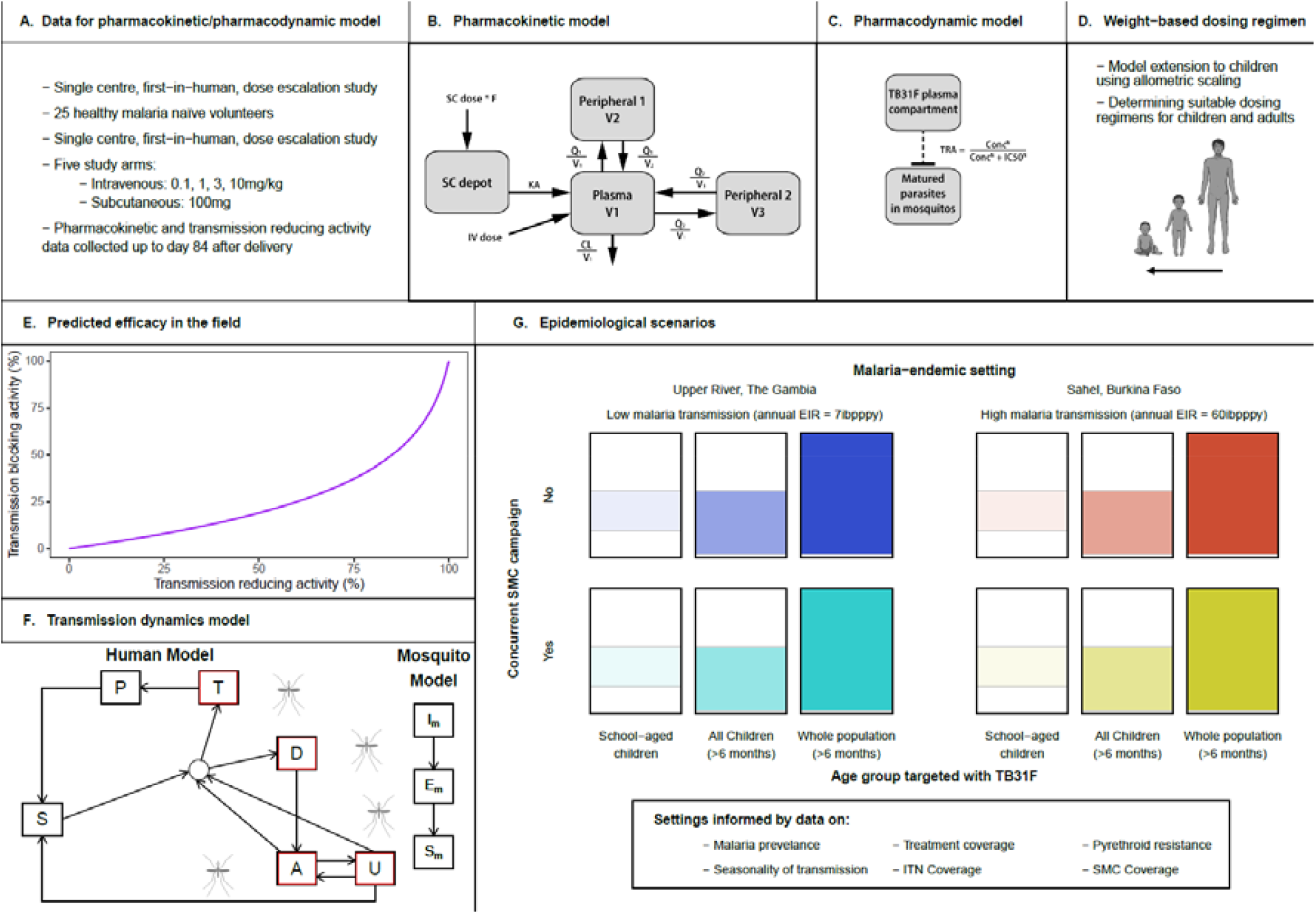
Overview of the pharmaco-epidemiological modelling workflow. **(A)** We used data from the first-in-human trial of TB31F (15) for pharmacokinetic-pharmacodynamic modelling. **(B)** The final pharmacokinetic model consisted of three disposition compartments and an absorption compartment for subcutaneously administered doses. **(C)** The pharmacodynamic model described the relationship between TB31F concentrations and transmission-reducing activity (TRA) using an Emax model. **(D)** By extrapolating the pharmacokinetic model from adults to children by allometric scaling, we proposed a weight-based dosing regimen that is predicted to obtain a similar duration of effective TRA in children >6 months of age compared to adults. **(E)** Predicted efficacy of TB31F in blocking malaria transmission events in the field. We use previous modelling work (25) to generate an estimated transmission-blocking activity for a given TRA, informed by data on oocyst counts found in naturally infected, wild-caught mosquitoes from Burkina Faso. **(F)** Mathematical model of malaria transmission (23). Individuals transition between six states: malaria-susceptible (S), untreated clinical disease (D), treated clinical disease (T), asymptomatic microscopy-detectable infection (A), asymptomatic submicroscopic infection (U). Infected humans (outlined in red) can transmit malaria to mosquitoes. Mosquitoes can be susceptible to malaria (*S*_*M*_), infected but not yet infectious (*E*_*M*_), or infectious (*I*_*M*_). **(G)** Modelled settings for the introduction of TB31F. We modelled the introduction of TB31F in two different seasonal malaria settings: a high-transmission site, based on the Sahel region of Burkina Faso, and a low-transmission site, based on the Upper River region of The Gambia. The values of the EIR shown are baseline values (i.e. no interventions against malaria implemented). We vary the age group targeted with TB31F (the coloured area of each rectangle indicates the proportion of the community targeted), administering the mAb to school-aged children (5-15 years of age), all children over 6 months (0.5-15 years of age), and the whole community (excluding children under 6 months of age). The intervention is delivered alongside (or instead of) seasonal malaria chemoprevention for children aged 3-59 months. SC, subcutaneous; IV, intravenous; KA, absorption constant, V, volume; Q, inter-compartmental clearance; CL, clearance. TRA, transmission-reducing activity; CONC, TB31F concentration in the central pharmacokinetic compartment; N, Hill factor; IC50, TB31F concentration obtaining 50% TRA; SMC, seasonal malaria chemoprevention; EIR, entomological inoculation rate; ibpppy, infectious bites per person per year; ITN, insecticide-treated bed nets; mAb, monoclonal antibody.

In previous work (25), a model of a hypothetical transmission-blocking vaccine was added to the transmission model to assess the potential impact such a vaccine could have and to identify key age groups to target for vaccination and the mathematical relationship between TRA and TBA (25). We utilise this relationship here, and also perform a sensitivity analysis to assess how it influences the public health impact of a transmission-blocking intervention. Unlike the previous work, the modelling framework used here allows for inter-individual variation in the pharmacokinetic model, as well as weight-dependent dosing. In the transmission model, the age of individuals is tracked, rather than their weight, we therefore utilised a weight-for-age model (adapted from Wasmann et al. (26), see Supplementary Methods) to enable a weight-based dosing regimen to be used. Within the transmission model, the TB31F concentration is described over time, using the population pharmacokinetic model described in Supplementary Table 1, for those individuals targeted with the intervention. On a given day, an individual’s TB31F concentration determines the reduction in the human to mosquito transmission probability.

We investigated the impact of introduction of TB31F in African settings with seasonal malaria transmission, contrasting a low (Upper River Region in The Gambia) and high endemic site (sahel region in Burkina Faso). In the supplemental information, we report details on seasonality (based on rainfall patterns (27)), transmission intensity (based on *P. falciparum* parasite prevalence in 2-10 year olds (PfPR_2-10_) (28)) and coverage and efficacy of ITNs, antimalarial treatment coverage (28) and SMC (1). With these interventions in place, we varied the mosquito-to-human ratio in the model, to obtain the desired level of endemicity. In the Sahel region of Burkina Faso, the mean PfPR2-10 in 2019 was 20%, with parts of the region having a PfPR_2-10_ as high as 30%. We elected to model a site with a relatively high prevalence for the region: therefore, we set a baseline EIR of 60 infectious bites per person per year (ibpppy), which results in a modelled PfPR2-10 of 29%. The observed PfPR2-10 in our second scenario, the Upper River region of the Gambia, was much lower (average value across the region in 2019 was 6%). We matched this observed prevalence using a baseline EIR of 7 ibpppy in the model.

We assumed that TB31F is administered annually, concurrent with the first SMC dose: in each setting this time point was chosen to optimise SMC efficacy relative to the seasonality of the region. In our default scenario, against which the impact of SMC and TB31F are measured, SMC is withdrawn for a three-year period, leaving ITNs as the only public health intervention (other than treatment for symptomatic malaria). We measured the impact of SMC and TB31F, used separately and in combination (Figures 3d and 4d), in terms of the number of clinical cases prevented over this three-year period (years 0 to 3 in Figures 3b and 4b). A three-year period was selected as this is the frequency of mass ITN campaigns in the region. We showed how the impact of administering the TB31F changes depending on the age group targeted: school-aged children (5-15 years of age), all children (up to 15 years of age, excluding infants under 6 months of age), and the whole community (excluding infants under 6 months of age). For all scenarios, we assumed a coverage level of 80% in the targeted age group for both SMC and TB31F. For the simulations of the high-transmission site, we simulated a population of 25,000. For the low-transmission site, we increased the population size to 50,000, as the lower incidence of malaria led to larger between-simulation variation. For, the public health projections, we ran 50 simulations and stated the mean impact of TB31F and SMC.

## Results

A visualization of our workflow is shown in Figure 1. We used data from the first-in-human TB31F trial (panel A) to develop a model describing its pharmacokinetics (panel B). Following the pharmacokinetic model, a pharmacodynamic model was developed describing the concentration-dependent TRA of TB31F, i.e. the reduction in mosquito oocyst burden related to mAb concentration (panel C). The pharmacokinetic model was extended to children and a dosing regimen that obtains effective exposure over a period covering the malaria transmission season for most Sub-Sahara African settings in both children and adults was identified using simulations (panel D). TBA, i.e. the ability of TB31F to reduce the proportion of mosquitoes that become infected, was calculated for simulated individuals over time using the predicted TRA (panel E). The time-varying individual-level TBA was used in a transmission model to explore the impact of different implementation strategies with TB31F on clinical malaria cases in twelve different epidemiological scenarios (panels F & G).

### Pharmacokinetic-pharmacodynamic modelling

Data from 275 pharmacokinetic samples of TB31F (from 24 individuals) and 3358 dissected mosquitoes from SMFA experiments were used in the pharmacokinetic-pharmacodynamic analyses. The final pharmacokinetic-pharmacodynamic model schematic is shown in panel 2 of Figure 1 and the final model parameters and goodness-of-fit plots are included in the online supplementary material. TB31F pharmacokinetics were best described by a linear model with three disposition compartments and an absorption compartment for subcutaneous doses. The subcutaneous bioavailability fraction was estimated at 0.54 (95%CI: 0.45-0.67). The mean absorption time was estimated at 3.2 days (95%CI: 2.8-4.2 days) which is in line with previously reported values for subcutaneous injection of other mAbs (29). The relationship between TB31F concentration and TRA is shown in Figure 2a. The concentration achieving 80% TRA was estimated at 3.43 mg/L (95%CI: 3.34-3.53 mg/L). This level of TRA, historically used as threshold for potency of transmission-blocking vaccines (30), was used for dose selection. Given available data on the safety and efficacy of intravenous administration (15), we used this route of administration for our further analyses. We extrapolated the pharmacokinetic model to children and explored weight-based dosing, aiming to reach an equivalent duration >80% TRA in children as observed with 10 mg/kg in adults. With a single dose of 100 mg for individuals with weight ≤10 kg, 300 mg for individuals weighing 10-15 kg and 700 mg for individuals >15 kg, the median duration over which a mAb concentration associated with >80% TRA was sustained was >5 months. (Figure 2b). Using this dosing regimen, this effective duration was comparable across all weights (Figure 2c) and was thus used in the simulations described from here on.

**Figure 2.**
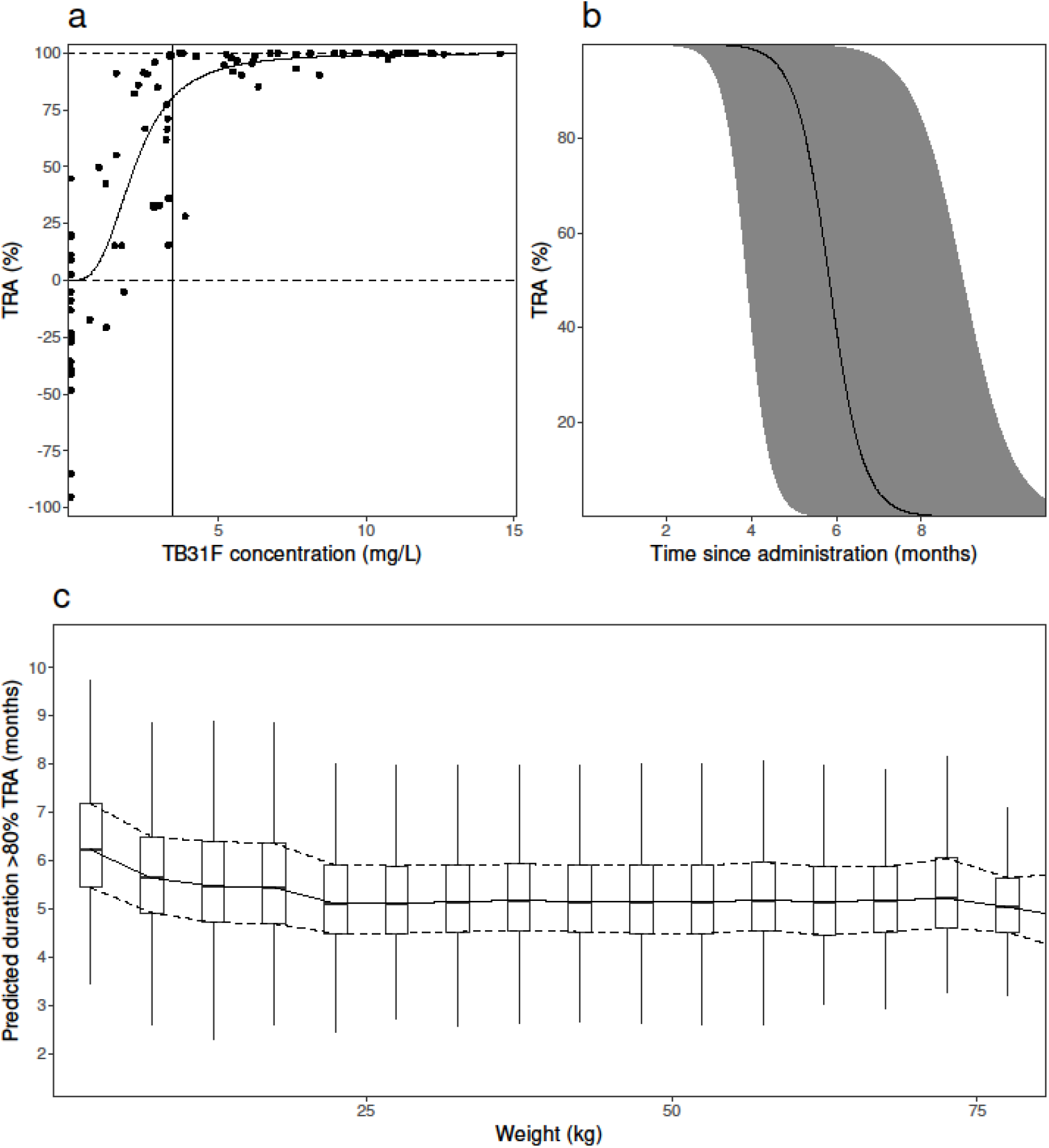
**(a)** Model-predicted relationship between TB31F concentration and transmission-reducing activity (TRA). The solid line represents the model-predicted TRA and the dots represent observed TRA. The vertical line indicates the concentration resulting in 80% TRA (a TB31F concentration of 3.43 mg/L; 95%CI: 3.34-3.53 mg/L). **(b)** Simulated TRA over time for all ages. The dose administered is 100 mg intravenously for individuals weighing under 10 kg; 300 mg for 10-15 kg; 700 mg for >15 kg. The continuous line represents the median and the band the 95% prediction interval. This regimen was used for in the following transmission modelling. **(c)** The duration >80% TRA is predicted to be similar across all weights using the proposed dosing regimen. The boxes show the middle, upper and lower quartiles per weight band with a 5 kg width. The whiskers depict the lowest and highest value within 1.5 times the interquartile range of the lower and upper quartiles, respectively. The simulations for panels b and c were performed with n=114,000 virtual males and females of 6 months of age and older in equal proportions. Weights of these virtual individuals were simulated using a weight-for-age model appropriate for an African population. We used these demographics to extrapolate down to children 6 months of age with weights representative for their age, which is especially important as the allometric scaling of clearance based on weight is age dependent.

### Predicted public health impact

A standalone implementation of the population-pharmacokinetic model that accounted for inter-individual variation, a weight-for-age model, and weight-based dosing was developed (the R code will be added to the Supplementary Materials upon publication). This model was incorporated into an established mathematical model *of Plasmodium falciparum* malaria transmission, in order to predict the impact that TB31F could have in two settings where malaria transmission is highly seasonal. These settings were tailored to resemble malaria transmission in the Sahel region in Burkina Faso, where malaria transmission is high (Figure 3), and the Upper River region of the Gambia, where malaria transmission is considerably lower (Figure 4). In both settings, insecticide-treated nets (ITNs) are distributed every three years, and children aged between 3 and 59 months receive seasonal malaria chemoprevention (SMC) during the transmission season (Figure 3a, Figure 4a). We simulated the impact of annual administration of TB31F, with a coverage of 80% of the target age group, for three consecutive years. This period was chosen to match the frequency of ITN campaigns. To enable a comparison between SMC and TB31F, SMC was withdrawn in our baseline scenario, leaving ITNs as the only active public health intervention. Although we are not proposing that TB31F could replace SMC in malaria-endemic settings, withdrawing SMC in our simulations enables a comparison of the community-wide impact of these interventions to be made. We modelled the administration of TB31F to three different target populations: school-aged children (aged 5-15 years), all children excluding young infants (6 months-15 years) and the entire population (excluding children under 6 months of age). For all simulations, we assumed 80% intervention coverage in the target population.

**Figure 3.**
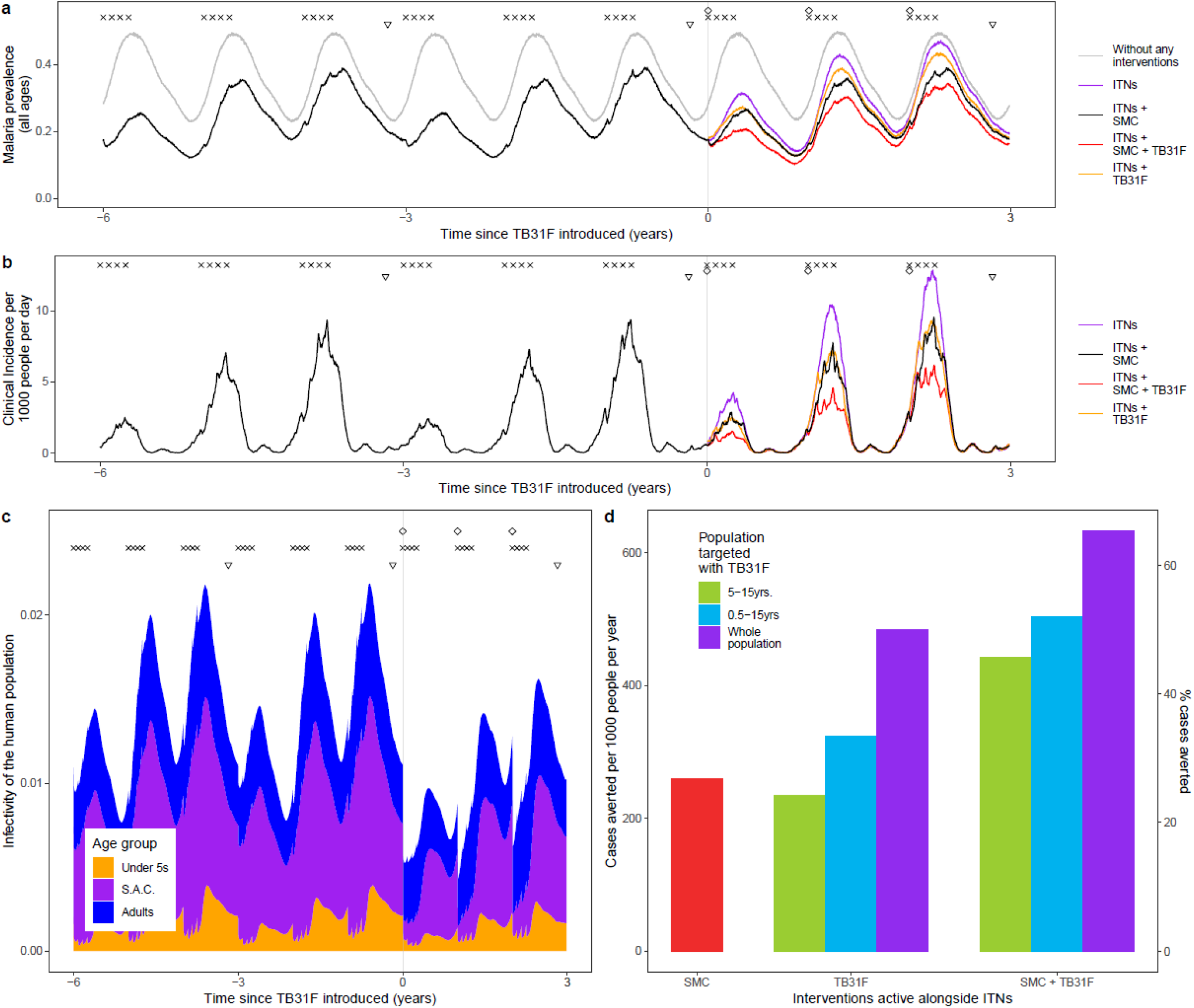
Modelling the introduction of TB31F intervention in the Sahel region of Burkina Faso (high-transmission setting). **a)** Malaria parasite prevalence (by microscopy) and changes over time as interventions additional to ITNs are introduced. The symbols at the top of the plot indicate the timings of intervention delivery. We assume that insecticide-treated net (ITN) distribution campaigns (triangles) and seasonal malaria chemoprevention (SMC) delivery (crosses) have occurred regularly prior to the introduction of TB31F (diamonds) at time zero. The grey curve indicates malaria prevalence prior to the introduction of control interventions. The coloured lines indicate the interventions used after time zero. SMC is delivered to children aged 3-59 months at 80% coverage; in panels a-c, TB31F is delivered to school-aged children (5 to 15 years of age) at 80% coverage. **b)** Model derived estimates of clinical incidence in the population over time. The impact of TB31F is most apparent in the second and third years of rollout, as the new ITNs prevent a high proportion of cases in the first year. **c)** The changing infectivity of the human population to a blood-feeding mosquito as interventions are introduced. This panel shows how the infectious reservoir (not adjusted for age-dependent mosquito biting rates) of malaria is distributed among three age groups: children under 5 years of age, school-aged children and adults (older than 15 years of age). After TB31F is introduced to school-aged children, the infectiousness of this age group is greatly reduced for several months. **d)** Cases averted due to SMC and TB31F interventions. Over a three-year period, we measure the reduction in cases on top of those prevented by ITNs. Here we vary the age group targeted with TB31F (school-aged children, all children over 6 months, and all age groups over 6 months in the community), assuming a coverage of 80% within the target age group. We quantify the clinical cases averted in two ways: the number of cases averted per 1000 people per year (left y-axis), and the percentage of the total number cases that is averted (right y-axis). In terms of cases averted per dose of TB31F administered, targeting children aged 5-15 years proved more efficient than targeting children aged 0.5-15 years, or the whole population over 6 months of age (see Supplementary Table 2).

**Figure 4.**
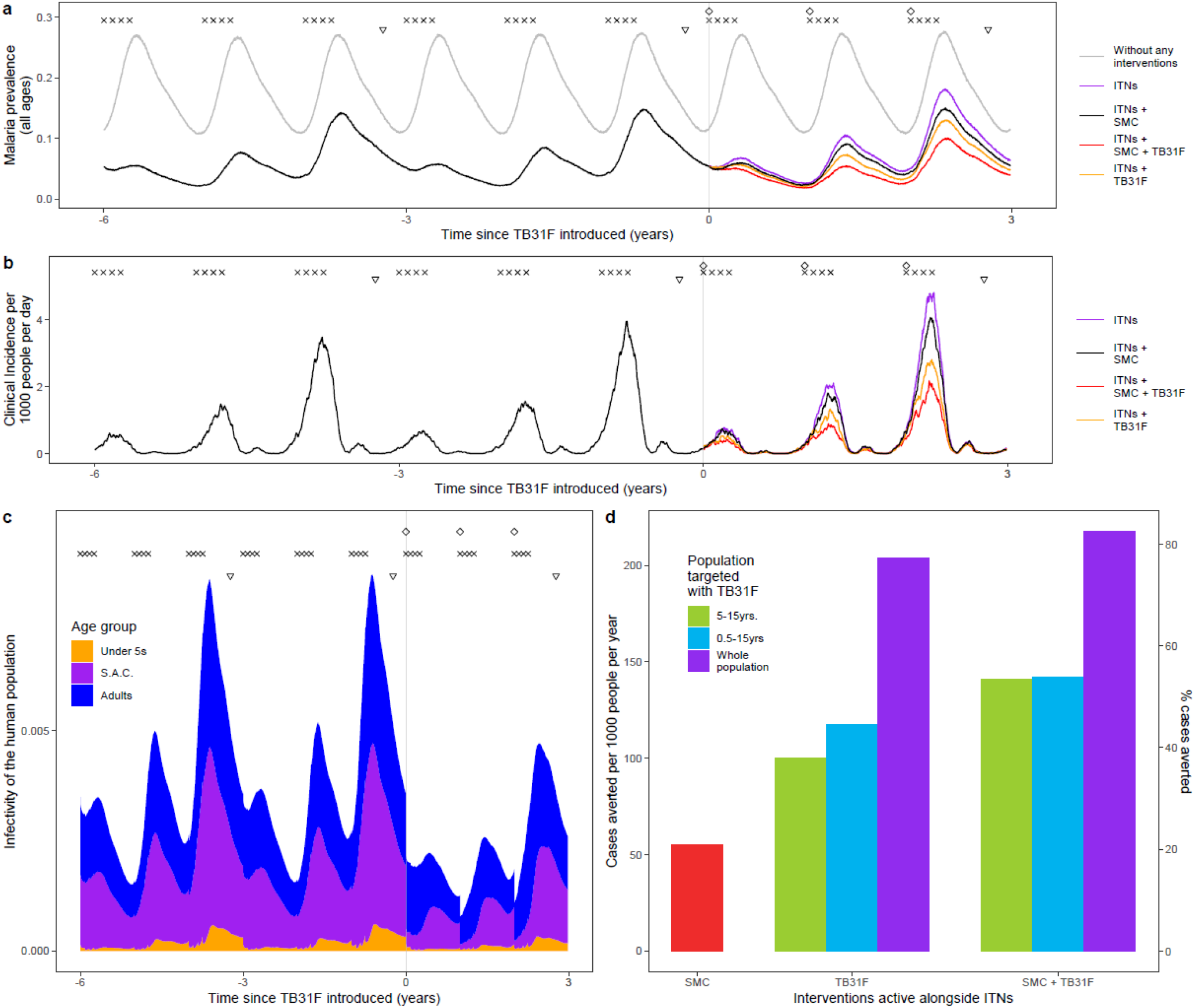
Modelling the introduction of TB31F intervention in the Upper River region of the Gambia (low-transmission setting). **a)** Here we show how modelled malaria parasite prevalence (by microscopy) is reduced as additional interventions to ITNs are introduced. The symbols at the top of the plot indicate the timings of intervention delivery. We assume that insecticide-treated net (ITN) distribution campaigns (triangles) and seasonal malaria chemoprevention (SMC) delivery (crosses) have occurred regularly prior to the introduction of TB31F (diamonds). The grey curve indicates malaria prevalence prior to the introduction of control interventions. ITNs, distributed every three years, have the largest impact on prevalence in the first year after distribution. SMC is delivered to children aged 3-59 months at 80% coverage; in panels a-b, TB31F is delivered to school-aged children (5 to 15 years of age) at 80% coverage. **b)** Modelled clinical incidence in the population over time. The impact of TB31F is most apparent in the second and third years of rollout, as the new ITNs prevent a high proportion of cases in the first year. **c)** The infectivity of the human population, as interventions are introduced. This panel shows how the infectious reservoir (not adjusted for age-dependent mosquito biting rates) of malaria is distributed among three age groups: children under 5 years of age, school-aged children and adults (older than 15 years of age). After TB31F is introduced to school-aged children, the infectiousness of this age group is greatly reduced for several months. **d)** Cases averted due to SMC and TB31F. Over a three-year period, we measure the reduction in cases on top of those prevented by ITNs. Here we vary the age group targeted with TB31F (school-aged children, all children over 6 months, and all age groups over 6 months in the community), assuming a coverage of 80% within the target age group. We quantify the clinical cases averted in two ways: the number of cases averted per 1000 people per year, and the percentage of the total number of clinical cases averted (right y-axis). In terms of cases averted per dose of TB31F administered, targeting school-aged children proved more efficient than targeting children aged 0.5-15 years, or the whole population over 6 months of age (see Supplementary Table 2).

In both malaria-endemic settings (Figures 3 and 4), TB31F was predicted to have a pronounced impact on clinical incidence across the community. In the high transmission setting, administering TB31F to children aged between 5 and 15 years has a similar impact compared to delivering SMC to young children (3-59 months). However, a much larger impact is seen when the interventions are delivered in tandem. In this region of Burkina Faso, delivering TB31F to school-aged children (aged between 5 and 15 years) in addition to delivering SMC to young children (3-59 months) is predicted to avert 48% of all clinical cases (462 cases per 1000 people per year) over a three-year period (Figure 3d), compared to the counterfactual scenario in which ITNs are the only active intervention. This value rises to 66% of cases (641 cases per 1000 people per year), if TB31F were administered to all age groups (Figure 3d). In the low transmission setting, administering TB31F to school-aged children had a larger impact than delivering SMC to children aged 3-59 months. As in the high transmission setting, delivering the two interventions in combination had a large impact: administering TB31F to school-aged children and also delivering SMC to children aged 3-59 months led to 53% of clinical cases (141 cases per 1000 people per year) being averted. This value rose to 82% (217 cases per 1000 people per year) when the TB31F campaign was extended to all age groups in the community. In both settings, targeting school-aged children with TB31F was the most efficient intervention in terms of cases averted per dose administered, but broadening the target population to also include other age groups substantially increased the overall impact of the intervention. The cases averted per dose of TB31F administered will vary depending on whether TB31F is delivered instead of or combined with SMC: we present the precise numbers in Supplementary Table 2. Our findings further suggest that, if coverage of SMC in children under 5 is high, there is little additional value in targeting this age group with a transmission-blocking intervention, especially in low transmission settings (Figure 4d, Supplementary Table 2).

The relative impact of targeting a specific demographic population is driven by the composition of the infectious reservoir, which changes when SMC and TB31F are introduced. Figures 3c and 4c show how the contribution of different populations to transmission changes over time when SMC and SMC plus TB31F are implemented; Figure 5 presents contributions to the infectious reservoir over the entire three-year period for the different age groups considered. In both the low (panels a-c) and high transmission (panels d-f), school-aged children (5 to 15 years of age) make an important contribution to the infectious reservoir (46% in the low transmission site, 49% in the high transmission site) prior to the implementation of SMC and TB31F. Compared to the high transmission setting, children under 5 years of age make a smaller contribution to the infectious reservoir in the low-transmission setting. When TB31F is administered to 80% of school-aged children alongside standard implementations of ITN and SMC, the overall infectious reservoir shrinks (by 26% in the low transmission site and 29% in the high transmission site) and the relative contribution of adults to onwards malaria transmission increases. In Supplementary Figure 6, we show the impact of delivering TB31F to 80% of the entire population in each setting (excluding children under 6 months of age), resulting in a more dramatic reduction in community-level infectivity.

**Figure 5.**
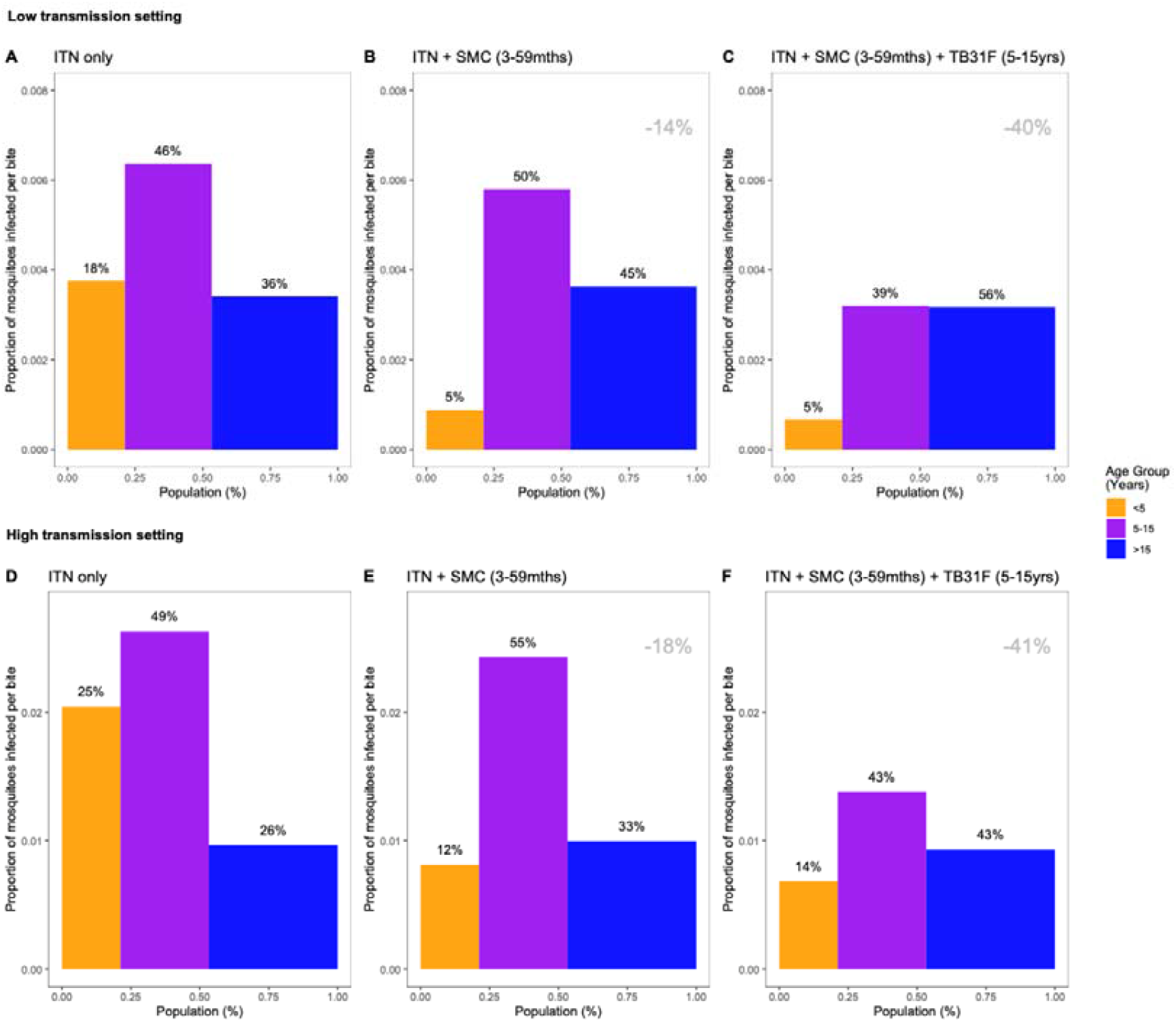
The changing infectious reservoir of malaria at the community level, as interventions are introduced. Model-derived projections of how different public health interventions against malaria influence the magnitude and composition of the human infectious reservoir. We show contributions to the infectious reservoir from three age groups: children under five years of age, school-aged children (5-15 years of age), and adults. These contributions are influenced by the average per-person infectivity (y-axes), and the relative sizes of the three subpopulations (x-axes). Panels A-C: modelling results from a low transmission setting, based on the Upper River region of the Gambia. Panels D-F: modelling results from a high transmission setting, based on the Sahel region of Burkina Faso. For both settings, we first assessed the infectious reservoir when the only intervention in use is insecticide-treated nets (panels A and D). We then assessed the impact of delivering seasonal malaria chemoprevention (SMC) to 80% of children aged 3-59 months (panels B and E). Thirdly, we measured the impact of delivering TB13F to 80% of school-aged children (panels C and F). In both settings, the introduction of TB31F reduces the infectivity in all age groups, due to a reduction in malaria transmission. It also results in a shift in the relative importance of age groups for the infectious reservoir, increasing the contribution that adults make to malaria transmission. The results in each panel were generated by averaging over a three-year period. Panels A and D correspond to a time-period prior to the introduction of SMC (not shown in Figures 4C and 3C). Panels B and E correspond to the three-year period (–3,0) in Figures 4C and 3C respectively; panels C and F correspond to the three-year period (0,3) in Figures 4C and 3C respectively. The percentage reductions (top-right corner of the middle and right panels) indicate the overall reduction in the infectious reservoir relative to the ITN-only scenario in each setting. To allow a direct comparison between settings we use a generic demography for sub-Saharan Africa (23). In Supplementary Figure 6, we repeat the analysis for the case where TB31F was administered to all age groups. Note that the results presented here are not adjusted for age-dependent biting of mosquitos.

An important factor in the predicted impact of a transmission-blocking intervention is the relationship between TRA and TBA (Methods). The relationship used here is informed by detailed data on naturally infected mosquitoes, collected in a high-transmission setting in Burkina Faso in 2014 (8). Parasite loads may vary between settings, e.g. due to transmission intensity, or vector composition. As detailed data such as those reported by Bompard et al. (8) are not often reported, we have used the same relationship between TBA and TRA for both sites examined here. In Supplementary Figure 7 we show the influence of parasite load on our modelling projections, by quartering and doubling the mean oocyst counts. We find that decreasing the mean oocyst counts does lead to a slightly larger public health impact, and *vice versa*. But our results are quite robust to changes in the TRA-TBA relationship, which has not been widely characterised *in natura*.

We also examined the potential impact of faster clearance of the mAb, which could be expected in the translation from the European trial participants to individuals living in malaria-endemic countries (Supplementary Figure 8). Studies reporting on the effect of cachexia describe a clearance that is around 1.33-fold higher (31–33). We find that a 1.33-fold increase in the clearance parameter in the pharmacokinetic model reduces the typical amount of time an individual maintains TRA>80% (Supplementary Figure 8A). This would reduce the public health impact of TB31F, but the modelling predicts that this reduction would be small (Supplementary Figures 8B & 8C). This is because the two settings selected here are highly seasonal: we expect that increasing the clearance would have a larger impact in settings with a longer transmission season, or malaria transmission throughout the year.

## Discussion

In this work, we have used an integrated approach that combined pharmacokinetic-pharmacodynamic modelling with an established malaria transmission model. We found that annual administration of a single weight-based dose at the start of the malaria season resulted in a substantial reduction in malaria transmission and clinical incidence. Of the three age groups we considered for TB31F administration, targeting school-aged children had the largest impact in terms of cases averted per dose administered. However, the total impact of the intervention was substantially increased when the other age groups were also targeted and when the mAb intervention was combined with SMC.

Interventions that specifically target the transmission of malaria differ from conventional malaria control measures by providing a delayed, rather than direct, benefit (9). Whilst transmission-blocking interventions benefit from clear and measurable biological efficacy endpoints in the reduction in mosquito infections, translating this efficacy to public health benefits is complex. In addition, the eligible population for transmission-blocking interventions is broader than that of, for example, SMC or the currently available RTS,S malaria vaccine that both primarily reduce malaria incidence in young children who are also the intervention recipients. Transmission-blocking interventions should be implemented in those populations that are (most) important for onward transmission. Importantly, the age groups that drive malaria transmission are not identical to those most at risk of severe disease (18,34,35). Recently, TB31F was tested in healthy adult volunteers where it had an excellent safety profile, including for the dose considered here, and post-infusion antibody kinetics and potency were indicative of prolonged transmission-blocking efficacy (15). However, antibody kinetics from adults cannot readily be translated to children (22). We translated TB31F’s pharmacokinetic properties in western healthy adult volunteers to a sub-Sahara African population of both children and adults by incorporating allometric scaling of pharmacokinetic parameters using body weight. We purposefully sought to identify a simple dosing regimen with wide age-bands receiving the same mAb dose to facilitate implementation of the intervention. With a simple strategy with three different doses, we predict >80% transmission reduction for a median duration of 5 months post intervention can be achieved. It should be noted that the 80% efficacy threshold has historically been considered for transmission-blocking vaccines (30), although lower efficacy levels can also have profound effects on malaria transmission (36).

We next simulated the introduction of TB31F into malaria-endemic settings in which ITNs and SMC for young children are well-established public health interventions. When targeting the entire population (excluding children under 6 months of age) with TB31F at 80% coverage on top of ITNs and SMC, a single annual administration reduced clinical malaria incidence by 54% and 74% over three years in high- and low-transmission settings, respectively. While the overall impact of TB31F will decrease when only certain age groups are targeted, the uneven contribution of different populations to onward transmission (18,34,37), allows operationally attractive demographic targeting to increase intervention efficiency. Choosing which age groups to prioritise with a transmission-blocking intervention relies on an understanding of who contributes most to malaria transmission in a particular setting. The infectious reservoir is shaped by a number of different mechanisms. In broad terms, long-lasting asymptomatic infections originate from two routes: symptomatic malaria infections that are not treated with drugs (or treated unsuccessfully), or infections that are sufficiently well controlled by the host’s immune response, due to naturally acquired immunity, so that symptoms do not develop. In untreated infections, asexual parasite density determines the density of gametocytes that is produced, which is a key influence in the probability that a feeding mosquito will become infected (38). Our work demonstrates that the impact of administering TB31F to a subset of the population, namely school-aged children (5-15 years of age), may achieve a comparable impact on clinical case load as SMC (given to children aged 3-59 months) in a high-transmission setting and that the impact may greatly exceed that of SMC in a low-transmission setting. Whilst we do not want to suggest that SMC may be withheld from populations in sub-Sahara African regions, where it achieves a very high level of personal protection against clinical malaria (39), our findings demonstrate the large public health impact a malaria transmission-blocking intervention can have. Our findings further indicate that combining SMC and TB31F may considerably increase the number of cases averted. Of course, TB31F may also be combined with other interventions. A recent clinical trial from Mali demonstrated that combining the pre-erythrocytic malaria vaccine RTS,S with SMC was superior to either intervention delivered alone (40). Our findings further highlight that interventions against malaria will alter the relative importance of different populations for the human infectious reservoir, suggesting that a periodic review of targeting strategies may be needed to maximize sustained intervention impact.

It is a significant advantage that only a single dose of TB31F is needed to achieve a prolonged duration of transmission reduction. This facilitates implementation on a large scale; a less potent mAb would require multiple doses per year or antibody half-life extension in order to achieve the same effective duration. A potent mAb that targets a protein on the surface of *Plasmodium* sporozoites was engineered to have about a 2-fold extended half-life (CIS43LS) and was recently demonstrated to induce strong anti-infection activity in human volunteers (41,42). Similar half-life extension technologies could also be of value for TB31F (42–44) and would lower the cost of the intervention by lowering the administered dose and may extend its effective period further. Even in areas of seasonal malaria, as examined in the current study, a non-negligible number of transmission events occurs outside the peak season and extending the effective duration following administration of TB31F would augment its public health impact (16). Half-life extension would be particularly beneficial when considering subcutaneous administration of TB31F, which is currently limited by a maximum volume that can be administered at a single injection site (100 mg was administered spread over two injection sites in the original trial). By extending the half-life, it would be possible to achieve >80% TRA for a longer duration with a similar injection volume. Although intravenous administration of a malaria vaccine has been achieved at a large scale (45), subcutaneous administration would definitely have considerable operational advantages.

Co-administration of a transmission-blocking intervention and pre-erythrocytic vaccine has already been shown to have a synergistic potential (46). Combining the anti-infection mAb CIS43LS with the transmission-blocking mAb TB31F also holds considerable promise. Not only would these two antibodies with distinctly different working mechanisms increase public health impact, TB31F may also prevent the spread of mutant parasites that escape CIS43LS. TB31F targets Pfs48/45, which has limited genetic variation, a shared advantage of many transmission-blocking vaccine candidates (11). The potency of TB31F has been demonstrated against two genetically distant parasite lines (15); the original rat mAb 85RF45.1 that formed the basis for TB31F has been tested against genetically diverse gametocyte isolates from Cameroon and Burkina Faso, further demonstrating universal activity and no indications for escape mutants (47).

Our study has a number of limitations. The pharmacokinetic extrapolation from healthy western adults to African populations has a solid mechanistic and empirical foundation(22,48), but dedicated studies of TB31F in African populations, including children, are needed to confirm its safety and the appropriateness of the proposed weight-based dosing regimen. These studies can confirm the pharmacokinetic properties in this population and test TB31F formulations to achieve the modelled doses by intravenous or subcutaneous administration. Differences in pharmacokinetic behaviour of monoclonal antibodies between healthy volunteers and the affected target population have previously been described, even in a western population (49). Cachexia, a type of disease-related malnutrition, is associated with a faster mAb clearance (32,50). Based on our sensitivity analysis (Supplementary Figure 8), we expect only a limited impact of the pharmacokinetic effects of cachexia, which may be more common in a malaria-endemic African population due to a higher incidence of malnutrition and comorbidities (51). With regard to our transmission model, it should be noted that the model contains no spatial structure and a mosquito is assumed not to preferentially return to feed on an individual or household it has already visited. This means the mAb-derived benefit is shared equally between those who received the intervention and those who did not. This may not always be the case: e.g. if certain households have very high intervention coverage and malaria transmission is highly focal. This could have implications for clinical trials of transmission-blocking interventions that measure epidemiological endpoints. Furthermore, the relationship between TRA and TBA depends on the oocyst intensity measured in the absence of the intervention (7). We have taken one relationship, generated from data collected in Burkina Faso in 2014, and applied it more widely. As this data was collected in a high-transmission setting, we expect that these oocyst intensities will be relatively high compared to many other settings. Our sensitivity analysis shows that the public health projections for the two settings considered here were fairly robust to changes to the TRA-TBA relationship. However, we expect the differences to become more apparent if TB31F were modelled in settings with a longer transmission season, or perennial malaria transmission. Finally, we have considered the deployment of TB31F in areas with low and high ongoing transmission and widespread use of ITNs and SMC. It may be that the preferred use case of mAb would be in lower transmission settings to try and push the disease to elimination. Predicting the epidemiological benefits of novel interventions in elimination settings is beyond the scope of this current work as it will depend on changing patterns of intervention use, the ability of local health systems to treat and diagnose cases, as well as the level of disease importation. These factors are currently relatively difficult to predict, though should be considered in further work.

## Conclusion

We have shown that the transmission-blocking mAb TB31F may be an effective intervention against malaria in settings with a well-defined malaria season. Targeting school-aged children of 5-15 years old showed to be most efficient for reducing malaria transmission and had a significant public health impact comparable to current implementations of SMC. If TB31F is implemented alongside SMC, we predict an even larger impact on the number of clinical malaria cases averted. Implementation of a transmission-blocking mAb alongside established interventions could play a crucial role in decreasing the global burden of malaria.

## Data Availability

The modelling code used to generate the results presented here will be made available upon publication

## Acknowledgments

We thank the participants and study personnel of the first-in-human trial. We are grateful to Dr. Elin Svensson from the Uppsala University and Radboudumc for her advice regarding the pharmacokinetic-pharmacodynamic modelling analysis of this study.

## Ethics Statement

All the data analysis conducted during this research was secondary and used studies that had obtained ethical approval previously from the appropriate organisations. The original study received approval from the Arnhem–Nijmegen Committee on Research Involving Human Subjects (NL69779.091.19). The study was done in accordance with the latest Fortaleza revision of the Declaration of Helsinki (2013), the Netherlands’ Medical Research Involving Human Subjects Act, ICH Good Clinical Practice standards, and local regulatory requirements

## Patient and Public Involvement

Patients or the public were not involved in the design, or conduct, or reporting, or dissemination plans of our research

## SUPPLEMENTARY MATERIALS

### Supplementary Methods

#### Pharmacokinetic/pharmacodynamic modelling

Parametric nonlinear mixed-effects modelling was performed using NONMEM version 7.4.1 to analyse the pharmacokinetic data and the relationship with transmission-reducing activity (21). Allometric scaling to a total body weight of 70 kg for volume, clearance and inter-compartmental flow parameters was included during parameter estimation from the start of model development to account for differences in weight, with exponents of respectively 1 and 0.75 for volume and clearance parameters. Different numbers of disposition compartments were explored to describe the pharmacokinetics of TB31F. Subcutaneous absorption was described using a dose depot and first-order absorption into the central compartment. Inter-individual variability was explored on the volume and clearance parameters of the pharmacokinetic model.

The pharmacodynamic model was developed following the pharmacokinetic model, using individual pharmacokinetic parameter estimates as input. The relationship between TB31F concentrations and transmission-reducing activity was modelled using oocyst count data of individual mosquitos as analysed in the standard membrane feeding assay (SMFA). In the SMFA, the number of oocysts per mosquito was determined for 20 mosquitos per pharmacokinetic blood sample The distribution of oocyst counts was described by a negative binomial distribution function for which mean baseline oocyst count and overdispersion were estimated. Inter-assay variability was included on the mean baseline oocyst count. An Emax model was used to characterise the relationship between TB31F concentrations and reduction in oocyst count (TRA) using the following equation:

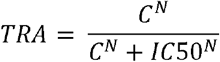

Where TRA is the proportion reducing the mean baseline count of the negative binomial distribution, C is the TB31F concentration in the central pharmacokinetic compartment, N is the Hill factor and IC50 is the concentration at which 50% effect is observed.

The pharmacokinetic model parameters were estimated using the first-order conditional estimation method with interaction and the pharmacodynamic parameters using the Laplacian estimation method. The inter-individual and inter-assay parameters in the pharmacokinetic and pharmacodynamic models were assumed to be log-normally distributed. Residual variability in TB31F pharmacokinetics was implemented using a proportional error model. The uncertainty of the pharmacokinetic and pharmacodynamic parameters was determined using sampling importance resampling (52). Model selection and evaluation was based on physiological plausibility, goodness-of-fit plots and comparison of the objective function value. A change in objective function value of 3.84 points between two nested models, associated with a significance level of 0.05, was considered statistically significant. Non-nested models were compared using the Akaike Information Criterion.

We developed a practical weight-based intravenous dosing regimen reaching an equivalent duration >80% TRA in children as observed with the highest dose tested in adults using simulations with the pharmacokinetic/pharmacodynamic model. The pharmacokinetic/pharmacodynamic model was extended from adults to children by implementing an age-dependent exponent for allometric scaling of pharmacokinetic clearance parameters (22). An exponent of 0.75 was used from the age of 5 years onwards. For younger children 2 to 5 years and 6 months to 2 years of age, exponents of 0.9 and 1.0 were used, respectively. We simulated female and male age groups of 6 months and every year from 1 to 18 years. Every group included 5000 virtual individuals. Weight data was simulated as described in a separate Supplemental Methods section. Weight-based dosing regimens were explored aiming to reach a time to 80% transmission-reducing effect similar to a 70 kg adult administered 10 mg/kg TB31F intravenously.

#### Weight-for-age modelling methods

Weight data was simulated following a previously described method using LMS data (26). This method uses reference values for weight by sex and age for children provided by the World Health Organization and Centers for Disease Control and Prevention growth charts (56,57). These growth charts allow calculation of the individual weight-for-age z-score, a measure of how many standard deviations the weight of a child differs from the median given their sex and age. This score is determined by sex- and age-specific parameters for the Box-Cox transformation (L), median (M) and coefficient of variation (S) which are reported in the growth charts. The z-score comes from a normal distribution with a mean of zero and standard deviation of one. Following this distribution, the weight of children may also be simulated using these charts. While these growth charts describe optimal development of children, Wasmann et al. additionally describe a correction accounting for stunted growth allowing more reliable simulation of an African population. A more detailed description of this workflow and the simulation script can be found in the original publication (26). As the charts stop at 18 years, simulation of adults was not incorporated in the workflow and we assumed the distribution at 18 years for all adults. This assumption will not hold in a western population, but is expected to be reliable in an African population.

#### Predicting public health impact using transmission modelling

The introduction of TB31F into two sites in west Africa is investigated, one of low malaria transmission, the other high. Using rainfall data, we chose two sites for which malaria transmission is highly seasonal. The high-transmission site was characterised to be representative of the Sahel region of Burkina Faso. For the rainfall patterns utilised for this region in the transmission model (27), 82% of the modelled annual clinical incidence of malaria occurs within 4 consecutive months. The low-transmission site was based on the Upper River region of the Gambia with 84% of the modelled annual clinical incidence of malaria occurs within 4 consecutive months. Before modelling the introduction of TB31F, we calibrated the model to reproduce the level of endemicity observed within each region according to the effectiveness of antimalarial interventions in use in the region. The level of endemicity was represented by estimates of the prevalence of falciparum malaria in 2-10 year olds (PfPR2-10) for the administrative unit 1 region from the Malaria Atlas Project (MAP) (28). To link transmission intensity to malaria prevalence, it is important to account for public health interventions currently in place. For these sites, we utilised data on ITN usage (60% in each setting, data from MAP (28)), treatment coverage for symptomatic malaria (50% in both settings, data from MAP (28)), and the coverage of SMC, which is administered as 4 rounds of sulfadoxine– pyrimethamine plus amodiaquine a year to children aged 3-59 months during the malaria season (80% in the Sahel region of Burkina Faso, 60% in the Upper River region of the Gambia, data from latest World Malaria Report (1)). For all MAP data utilised here, we obtained the latest estimates available through their website, as of January 2022 (all data pertain to 2019) (https://malariaatlas.org, accessed January 10^th^ 2022). The effectiveness of ITNs also depends on the levels of pyrethroid resistance observed in wild Anopheles mosquitoes in these regions (53), as this will inform the lethality of the ITNs used in the areas against the mosquitoes (54). In both regions it is assumed that all nets are pyrethroid-piperonyl butoxide ITNs and are distributed every three years in mass campaigns. For simplicity it is assumed each mass campaign achieves the same level of ITN usage (as defined above), after which usage and ITN effectiveness declines at realistic rates (54).

An important consideration in the modelling presented here is the relationship between TRA and TBA, which is governed by the parasite load in naturally infected, wild mosquitoes. The higher the parasite load, the harder it is for a transmission-blocking intervention to completely block the development of parasites within the mosquito (55). As outlined by Challenger et al. (25), we use a mathematical relationship between TRA and TBA. This relationship depends on the distribution of oocyst counts in naturally infected mosquitoes, which is described by a zero-truncated negative binomial distribution. The distribution is characterised by two parameters: m (which would be the mean of the distribution, were it not truncated) and k (the dispersion parameter). The relationship between TRA and TBA takes the following form (see Challenger et al. (25) for the derivation):

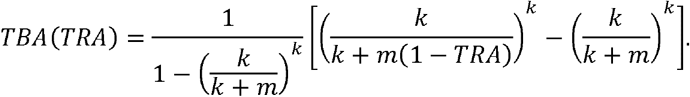

We used oocyst counts measured in naturally infected mosquitoes in Burkina Faso (8) to inform values of m and k Informed by the Burkina Faso data, we used parameter values m=0.000157 and k=0.00000495, which results in a mean oocyst count of 9.1. Note that here we fit one zero-truncated negative binomial distribution to the data: in Ref. (25) a more complex model, which assumes different TRA-TBA models for different malaria infections (e.g. symptomatic versus asymptomatic malaria), was also fitted. Here we choose the simpler approach. Data on oocyst counts in wild mosquitoes are rarely reported and may vary across settings. As a sensitivity analysis, we repeated the analyses used to generate Figures 3 and 4 whilst varying the mean oocyst count. We generated results for an adjusted TRA-TBA relationship, obtained by first quartering (m=0.0000158, mean=2.2), then doubling (m=0.000398, mean=18.2), the mean number of oocysts, holding the dispersion parameter fixed. The results of the sensitivity analysis are shown in Supplementary Figure 7, along with a plot of the modified TRA-TBA relationships.

**Supplementary table 1.** Final pharmacokinetic/pharmacodynamic model parameters.

| Model | Parameter | Estimate | 95%CI SIR |
| --- | --- | --- | --- |
| Pharmacokinetic | Central volume (L) | 2.57 | 2.19-3.00 |
|  | Peripheral volume 1 (L) | 0.94 | 0.61-1.27 |
|  | Peripheral volume 2 (L) | 1.47 | 1.17-1.78 |
|  | Clearance (L/h) | 0.0051 | 0.0045-0.0058 |
|  | Inter-compartmental clearance 1 (L/h) | 0.17 | 0.07-0.34 |
|  | Inter-compartmental clearance 2 (L/h) | 0.0078 | 0.0034-0.0164 |
|  | Subcutaneous bioavailability (fraction) | 0.54 | 0.45-0.67 |
| | Absorption rate constant ($\text{h}^{-1}$) | 0.013 | 0.010-0.015 |
|  | IIV clearance (%CV) | 30.0 | 22.9-41.2 |
|  | IIV central volume (%CV) | 35.2 | 27.0-49.0 |
|  | Correlation clearance-central volume (%) | 79 |  |
|  | Proportional residual error (%) | 18.5 | 16.9-20.0 |
| Pharmacodynamic | Baseline oocyst count (n) | 22.2 | 14.0-36.4 |
|  | Overdispersion parameter | 0.445 | 0.416-0.475 |
|  | IC50 (mg/L) | 2.18 | 2.12-2.24 |
|  | Hill factor | 3.05 | 2.94-3.15 |
|  | IEV baseline count (%CV) | 92.5 | 57.7-202 |

CI, confidence interval; CV, coefficient of variation; IIV, inter-individual variability; IC50, concentration at which 50% transmission-reducing activity is reached; IEV, inter-experiment variability; SIR, sampling importance resampling according to Dosne et al. (52). All pharmacokinetic parameters are allometrically scaled to a total body weight of 70 kg.

**Supplementary Table 2.**
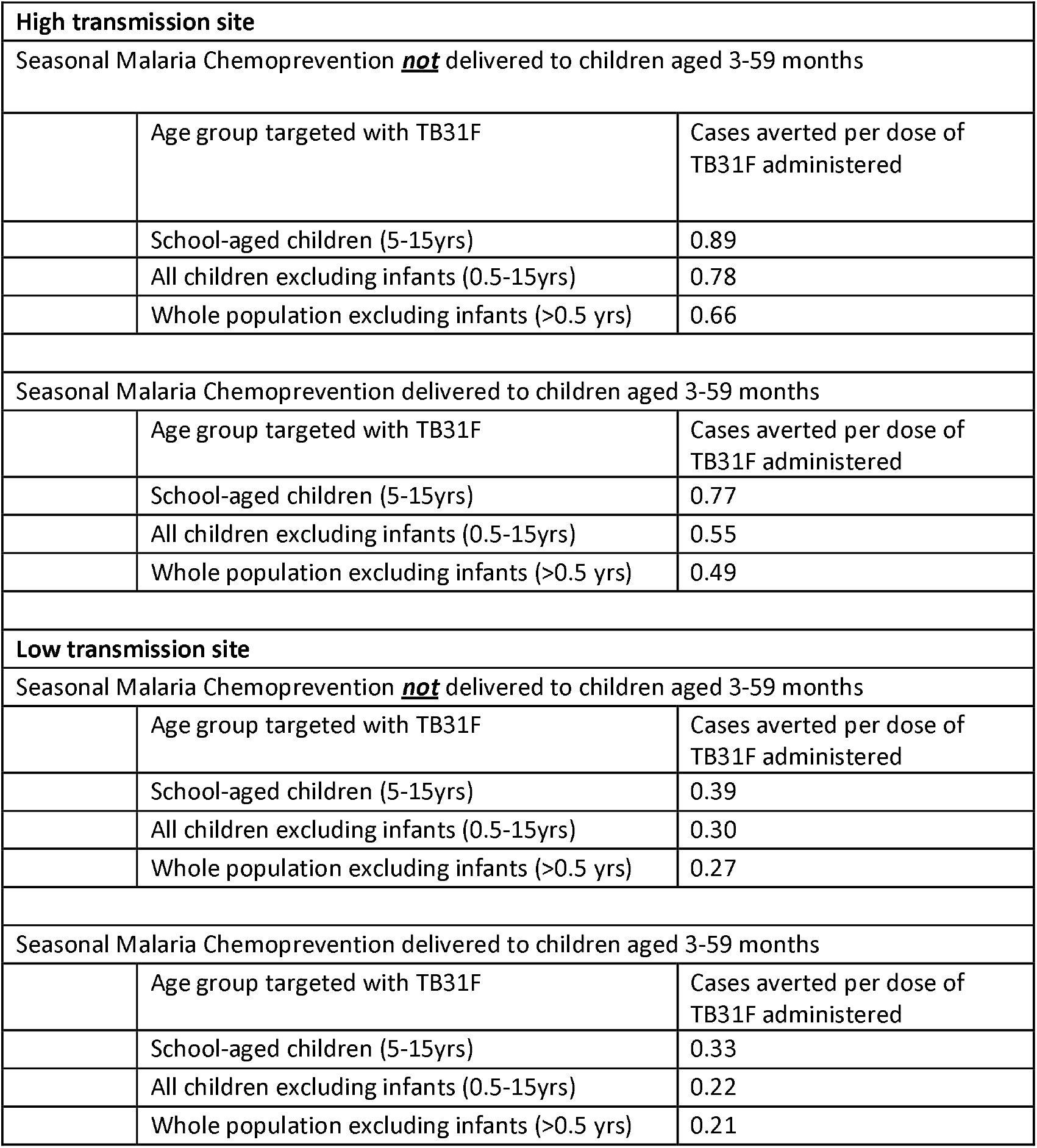
Cases averted per dose of TB31F administered. Here we report the clinical cases of malaria averted per dose of TB31F administered, for all modelling scenarios considered (as displayed in Figures 3 and 4). The impact of TB31F will depend on the age group targeted, as well as the other active public health interventions against malaria. In all scenarios, coverage of TB31F was 80% of the population targeted.

| **High transmission site** |  |  |
| --- | --- | --- |
| Seasonal Malaria Chemoprevention ***not*** delivered to children aged 3-59 months |  |  |
|  | Age group targeted with TB31F | Cases averted per dose of TB31F administered |
|  | School-aged children (5-15yrs) | 0.89 |
|  | All children excluding infants (0.5-15yrs) | 0.78 |
|  | Whole population excluding infants (>0.5 yrs) | 0.66 |
| Seasonal Malaria Chemoprevention delivered to children aged 3-59 months |  |  |
|  | Age group targeted with TB31F | Cases averted per dose of TB31F administered |
|  | School-aged children (5-15yrs) | 0.77 |
|  | All children excluding infants (0.5-15yrs) | 0.55 |
|  | Whole population excluding infants (>0.5 yrs) | 0.49 |
| **Low transmission site** |  |  |
| Seasonal Malaria Chemoprevention ***not*** delivered to children aged 3-59 months |  |  |
|  | Age group targeted with TB31F | Cases averted per dose of TB31F administered |
|  | School-aged children (5-15yrs) | 0.39 |
|  | All children excluding infants (0.5-15yrs) | 0.30 |
|  | Whole population excluding infants (>0.5 yrs) | 0.27 |
| Seasonal Malaria Chemoprevention delivered to children aged 3-59 months |  |  |
|  | Age group targeted with TB31F | Cases averted per dose of TB31F administered |
|  | School-aged children (5-15yrs) | 0.33 |
|  | All children excluding infants (0.5-15yrs) | 0.22 |
|  | Whole population excluding infants (>0.5 yrs) | 0.21 |

##### Pharmacokinetic/pharmacodynamic goodness-of-fit plots

**Supplementary Figure 1.**
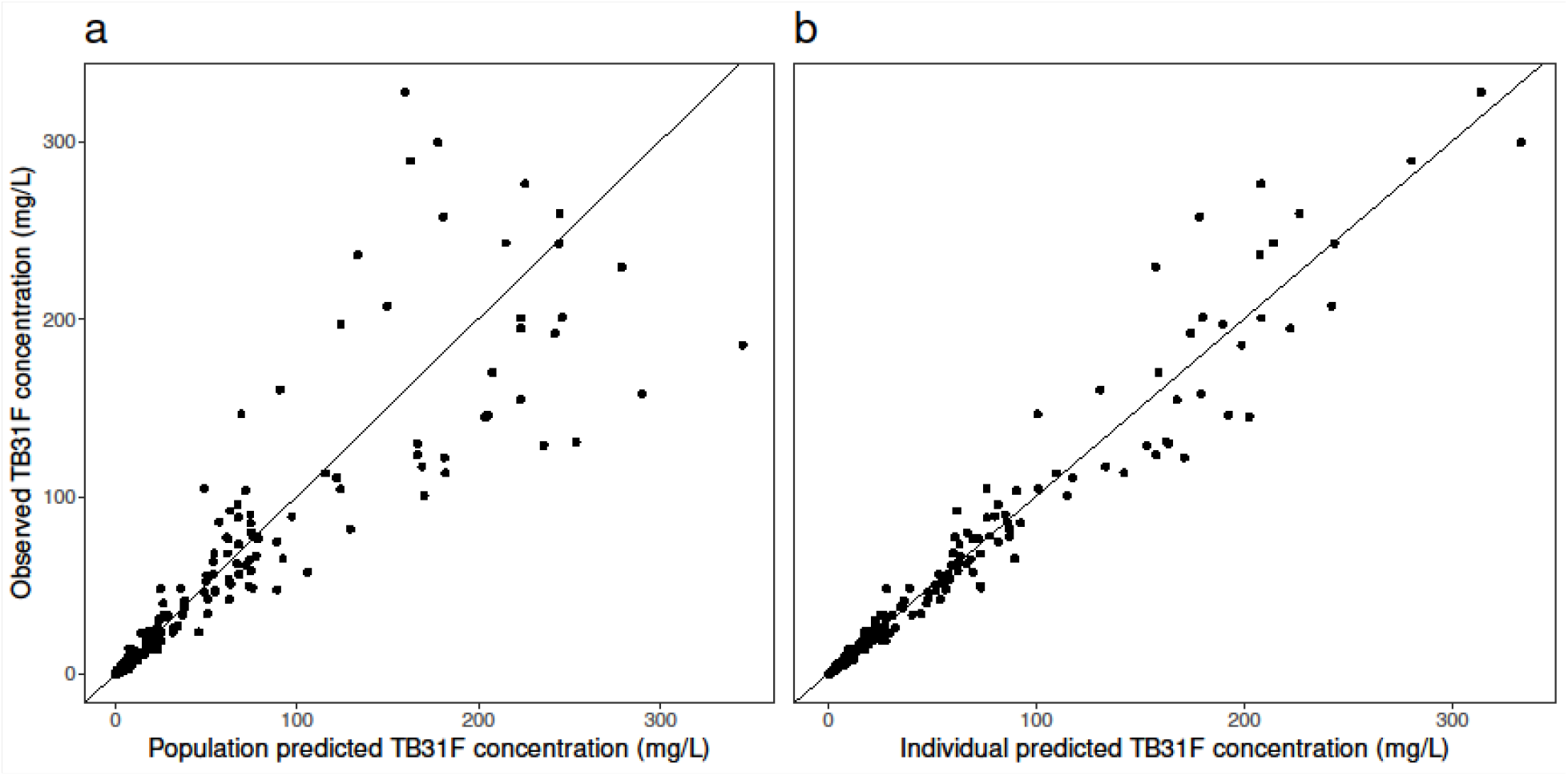
**(a)** Population and **(b)** individual model-predicted versus observed antibody concentrations. The solid black lines represent the line of unity.

**Supplementary Figure 2.**
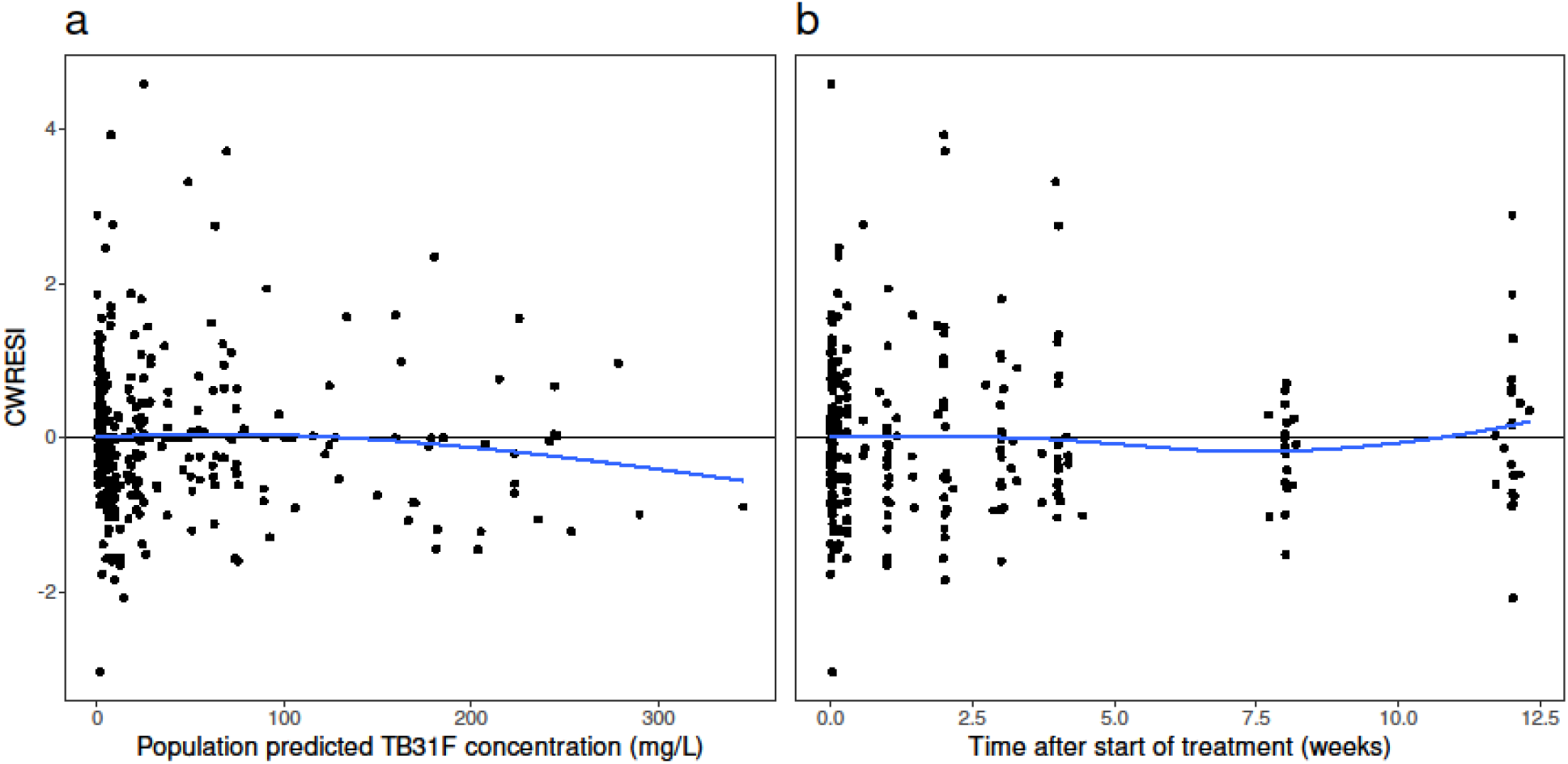
(**a**) Population model-predicted antibody concentration and (**b**) time after start of treatment versus conditional weighted residuals with interaction (CWRESI). The data points (dots) are evenly scattered around the zero-line (solid black lines) as shown by the smooth function (solid blue lines).

**Supplementary Figure 3.**
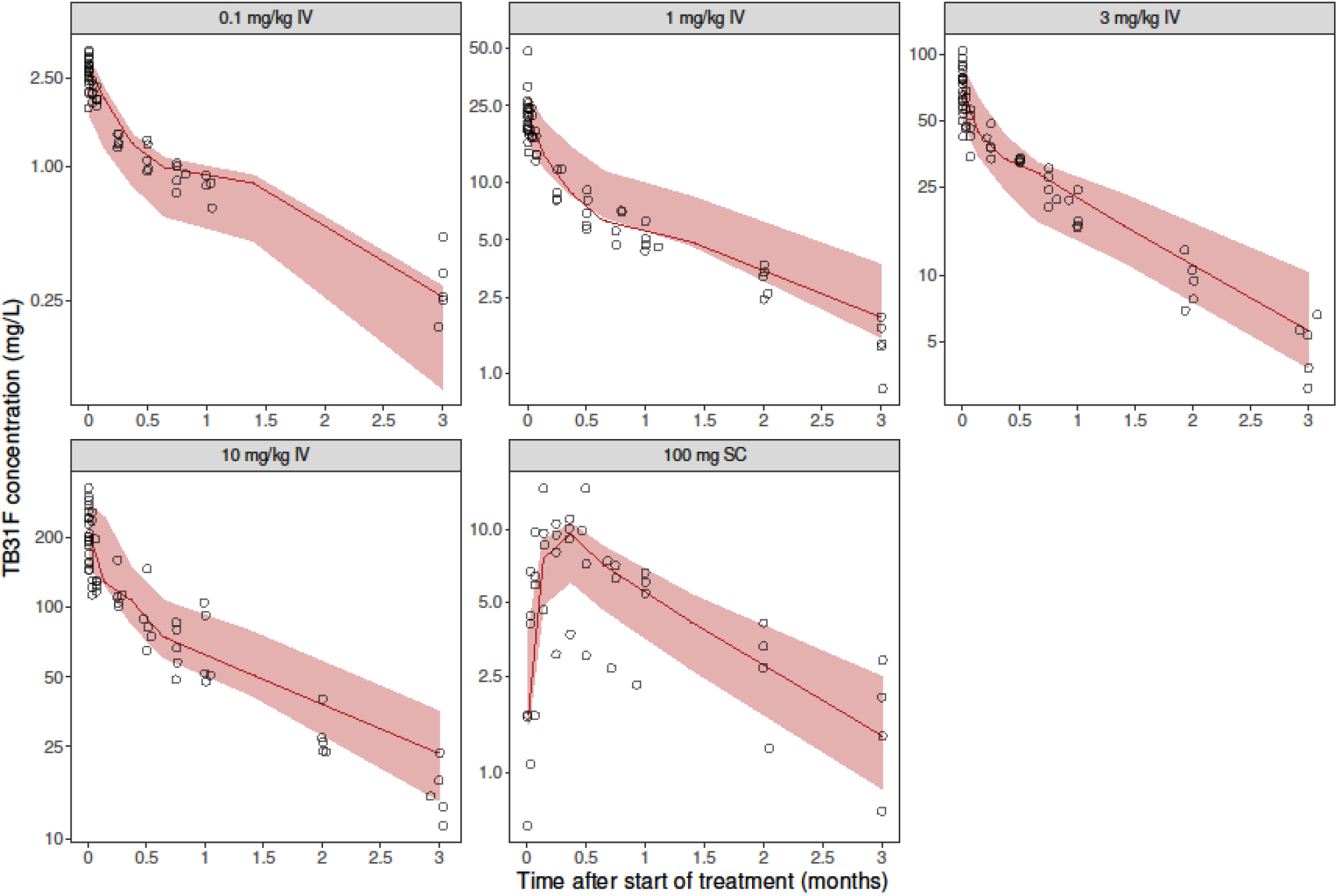
Observed and simulated TB31F concentrations over time per dosing group. The solid lines represent the 50^th^ percentiles of the observed TB31F concentrations depicted by the open circles. The shaded areas represent the simulation-based 95% confidence intervals of the 50^th^ percentiles based on n=1000 simulations.

**Supplementary Figure 4.**
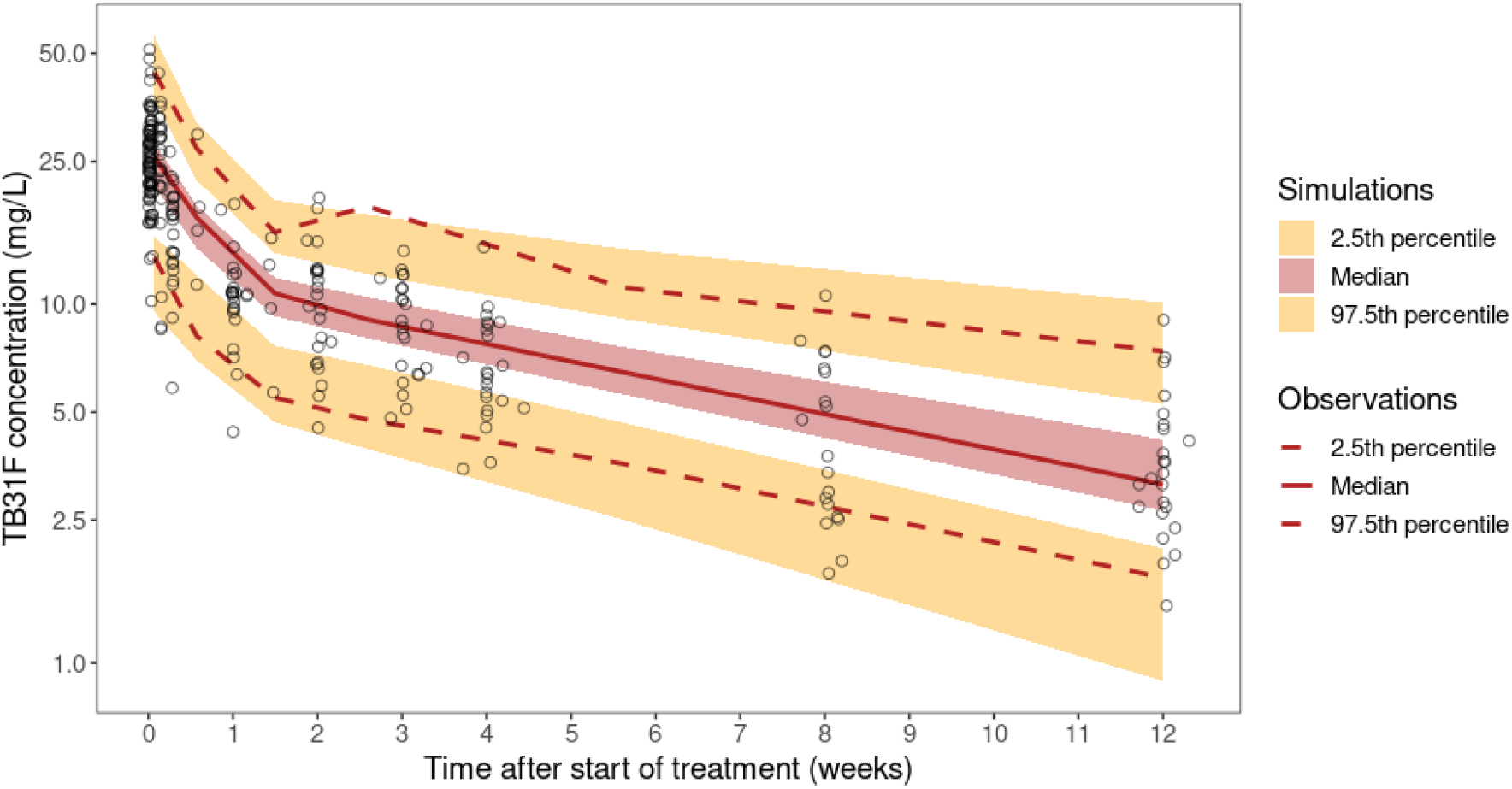
Prediction-corrected visual predictive check of the pharmacokinetic model based on n=1000 simulations. Observed and simulated TB31F concentrations are normalized to make them comparable between dosing groups (Bergstrand et al.). The solid line represents the 50^th^ percentile and the dashed lines represent the 2.5th and 97.5th percentiles of the observed TB31F concentrations depicted by the open circles. The shaded areas represent the simulation-based 95% confidence intervals of the 2.5th, 50th, and 97.5th percentiles. Bergstrand M, Hooker AC, Wallin JE, Karlsson MO. Prediction-corrected visual predictive checks for diagnosing nonlinear mixed-effects models. AAPS J. 2011 Jun;13(2):143-51.

**Supplementary Figure 5.**
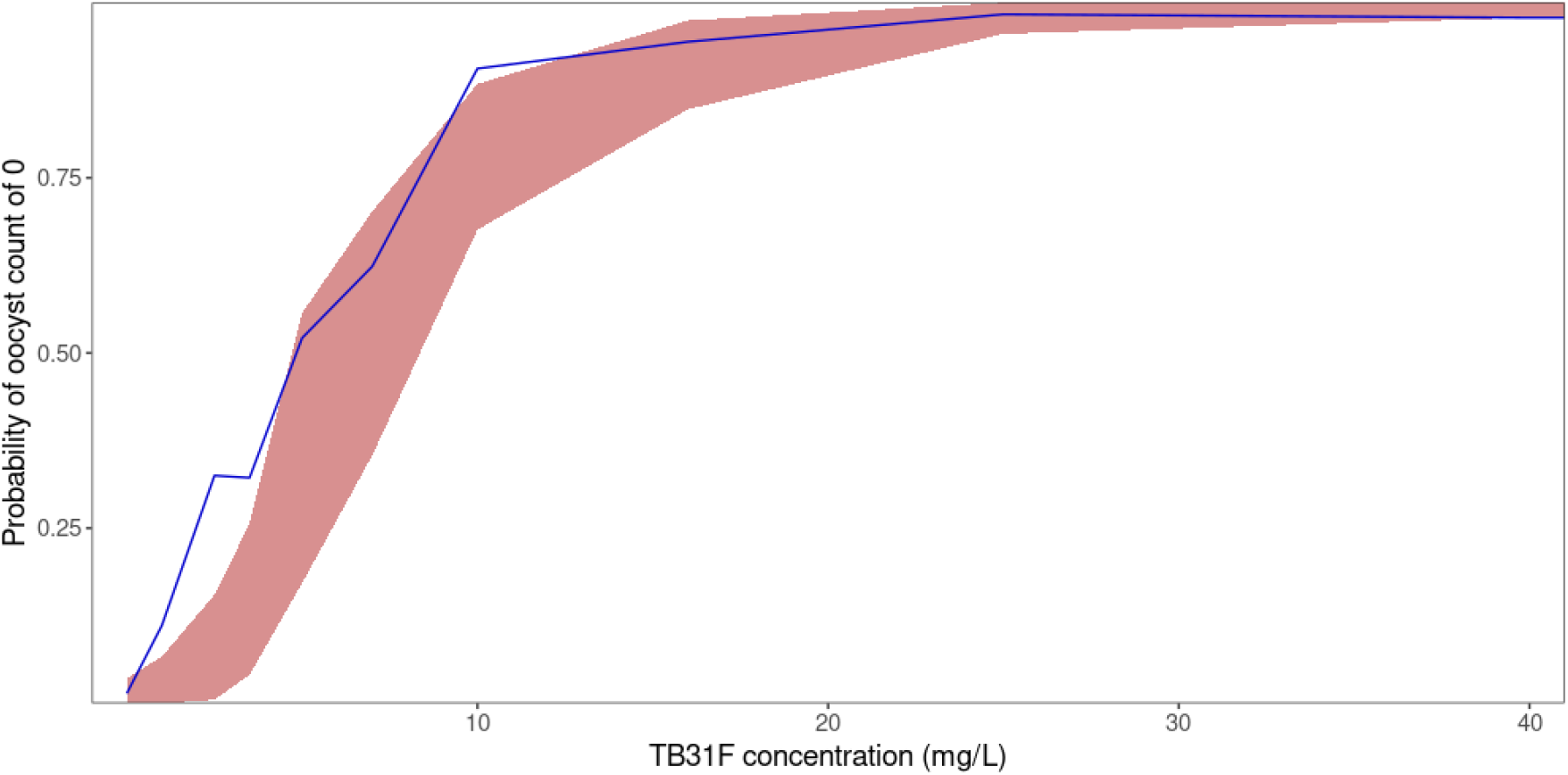
Observed and simulated antibody concentration versus the probability of an oocyst count of 0. The solid blue line represents the observed probability. The red band represents the 95% prediction interval of the simulated probability based on n=1000 simulations.

**Supplementary Figure 6.**
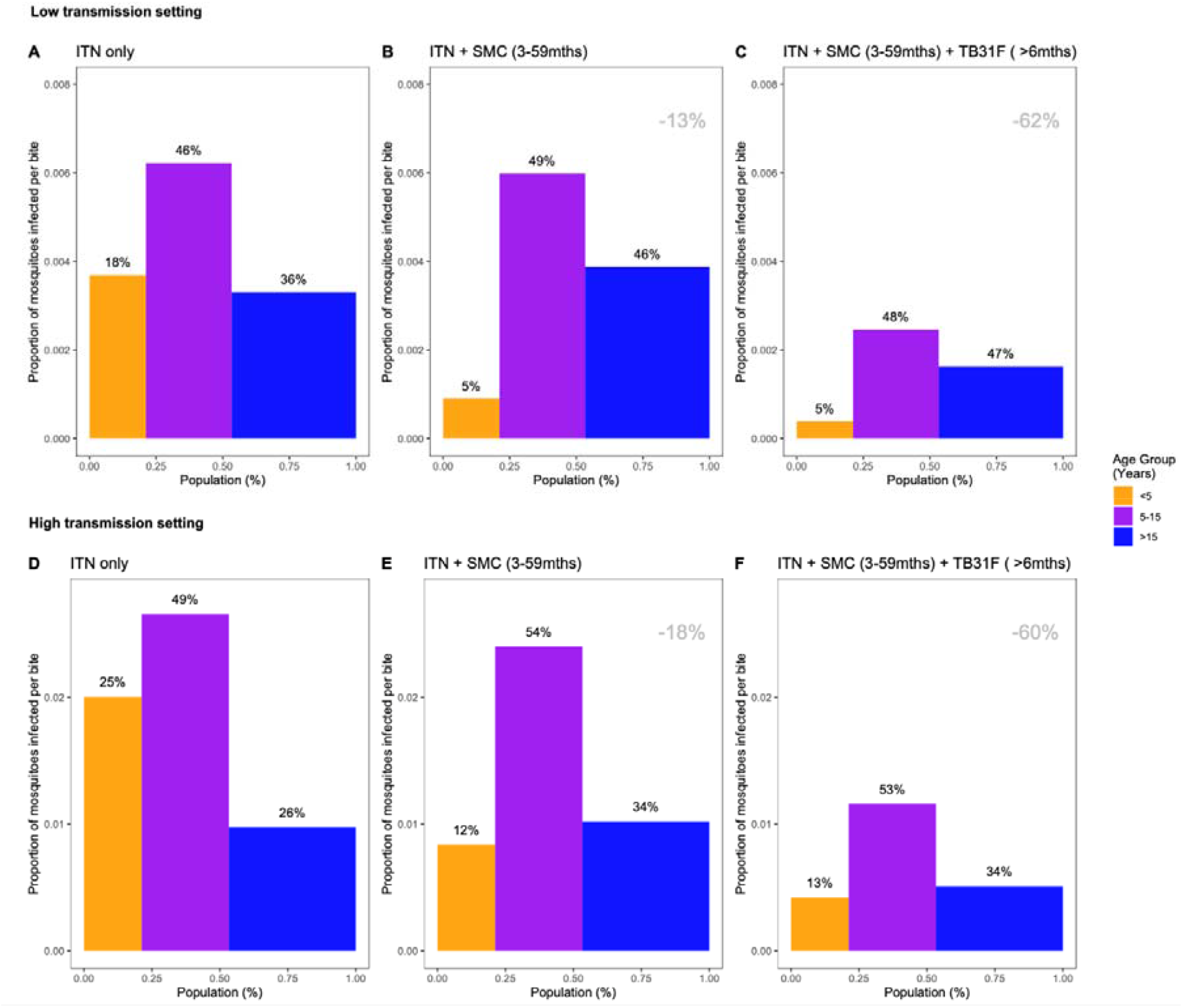
The changing infectious reservoir of malaria at the community level, as interventions are introduced (including the introduction of TB31F to all age groups). Here we show how the introduction of public health interventions against malaria can reduce the infectious reservoir at the community level. We show contributions to the infectious reservoir made by three age groups: children under five years of age, school-aged children, and adults. These contributions are influenced by the average per-person infectivity (y-axes), as well as relative sizes of the three subpopulations (x-axes). Panels A-C: modelling results from a low transmission setting, based upon the Upper River region of the Gambia. Panels D-F: modelling results from a high transmission setting, based upon the Sahel region of Burkina Faso. For both settings, we first assessed the infectious reservoir prior to the introduction of seasonal malaria chemoprevention (SMC) or TB31F, i.e. insecticide-treated nets (ITNs) are the only intervention in use (panels A and D). We then assessed the impact of delivering SMC to 80% of children between 3 and 59 months of age (panels B and E). Thirdly, we measure the impact of delivering TB13F to 80% of all age groups (panels C and F, excluding children under 6 months of age). The percentage reductions displayed in the top-right corner of the middle and right panels indicate the overall reduction in the infectious reservoir relative to the ITN-only scenario in each setting. For all results, a generic demography for sub-Saharan Africa was used, as per Griffin et al. (23). Note that the results presented here are not adjusted for age-dependent biting of mosquitoes.

**Supplementary Figure 7.**
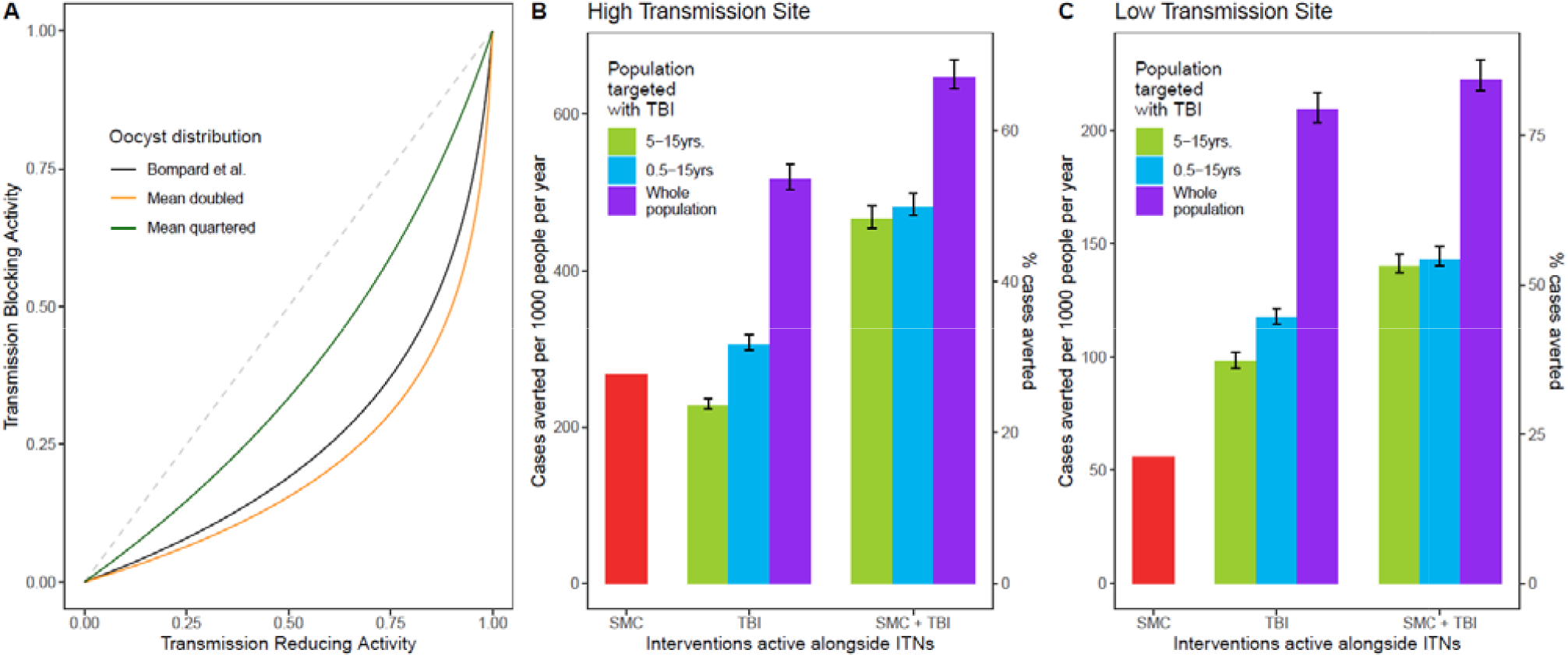
How changing the relationship between transmission-reducing activity and transmission-blocking activity affects the epidemiological impact of TB31F. In order to model TB31F as a public health intervention, we had to translate the measured pharmacodynamic properties of TB31F, calculated in terms of a transmission-reducing activity (TRA, Figure 2A), into a predicted efficacy for blocking human-to-vector transmission events in the field. This translation utilised data on oocyst counts in naturally infected, wild mosquitoes. This data was collected in a high transmission setting in Burkina Faso in 2014 (8), and was used to generate a relationship between TRA and transmission-blocking activity (TBA) in recent modelling work (25), which is shown by the black curve in panel A. This is the TRA-TBA relationship used throughout this work. The distribution of oocyst counts, which determines this relationship, is likely to vary across different malaria-endemic settings. Therefore, we varied the mean of this distribution (which is a zero-truncated negative binomial distribution), keeping the dispersion parameter fixed. The black curve is generated from a zero-truncated oocyst distribution, characterised by parameters *m*=0.000157 and *k*=0.00000495 (mean oocyst count=9.1). We double the mean of the distribution by setting *m*=0.000398, resulting in a TRA-TBA relationship shown by the orange curve in panel A. We quarter the mean, by setting *m*=0.0000158, resulting in a TRA-TBA relationship shown by the green curve in panel A. Panels B and C show the public health impact of the modelled administrations of TB31F, as described in Figures 3 and 4, in the high- and low-transmission settings, respectively. Decreasing the mean oocyst counts increases the impact of the public health interventions, and *vice versa* (changes indicated by the error bars). Therefore, the public health projections are robust to changes in the TRA-TBA relationship.

**Supplementary Figure 8.**
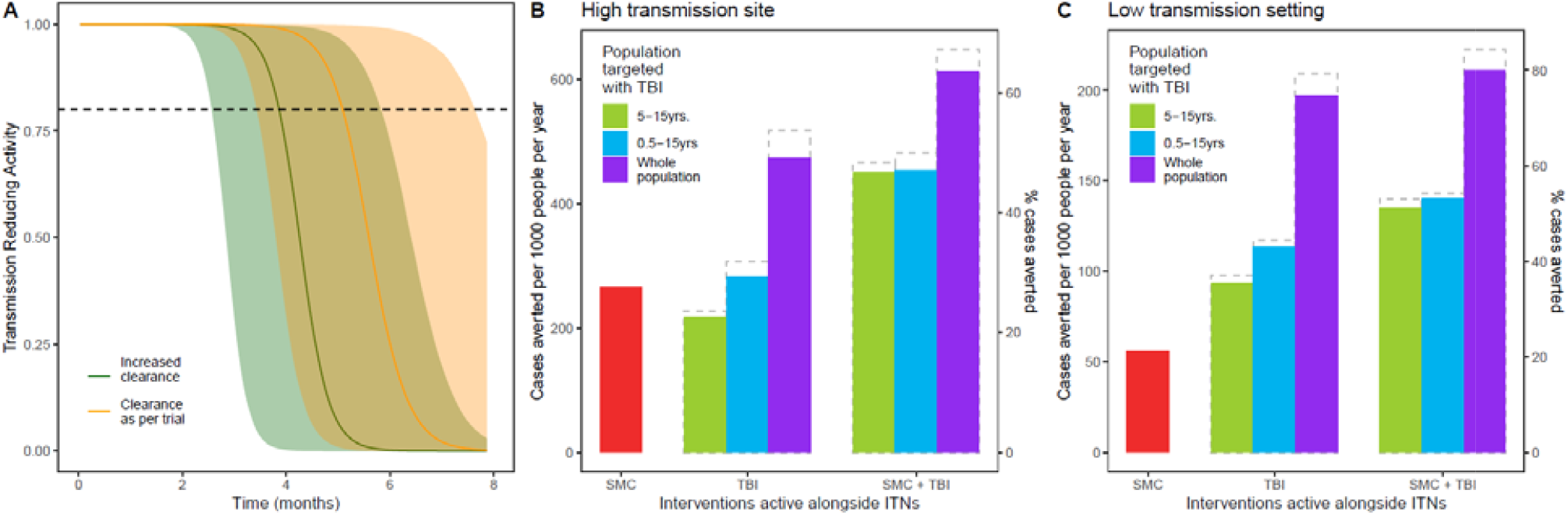
Impact of adjusting the clearance parameter in the pharmacokinetic model to account for a potentially faster clearance in the target population. In this sensitivity analysis, we explored the impact of a 1.33-fold increase in the population-average clearance parameter (that is, an increase from 0.0051 L/h to 0.0068 L/h) on the public health impact estimates of TB31F. Panel A: an increased clearance reduces the amount of time that that a transmission-reducing activity (TRA) of >80% can be maintained. These TRA projections were generated from PK modelling of two simulated cohorts, each of 4000 school-aged children. The age-distribution of these cohorts was consistent with that used in the transmission model (an exponential distribution with a mean of 21 years). Panels B and C: public health impact of increased clearance, in the high- and low-transmission settings, respectively. The modelling scenarios are as described in Figures 3 and 4. The original results, presented in Figures 3 and 4, are indicated by the grey, dashed bars. This indicates that the impact of TB31F would be slightly impacted by increased clearance in the cohort chosen for administration. One reason the impact is quite small is that the two settings chosen here are highly seasonal: this means that most clinical malaria occurs within the first 3 months after administration. During this time period, the impact of increased clearance upon the TRA of the modelled cohort is not so apparent: we expect a larger impact to appear, if TB31F was administered in a setting that was less seasonal.

**NONMEM control stream for the final pharmacokinetic model**

~~~
$PROB TB31F
$INPUT GROUP ID TIME AMT RATE DV EVID CMT BLQ SEX HT AGE WT
$DATA TB31F_NM.csv IGNORE=@
$SUBROUTINE ADVAN13 TOL=6
$MODEL NCOMPARTMENTS=4 COMP=(CENTRAL)
COMP=(PERI)
COMP=(DEPOT) COMP=(PERI2)
$PK
WTCL = (WT/70)**0.75
WTV = (WT/70)**1
~~~

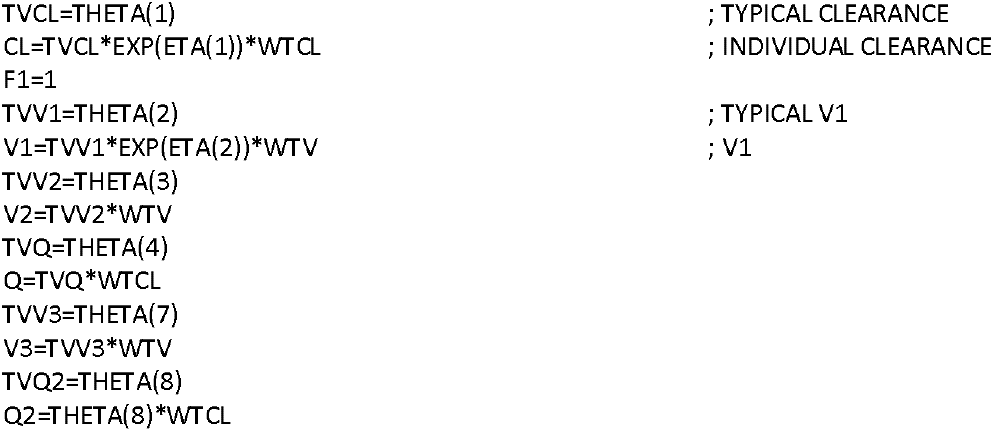

~~~
S1=V1
S2=V2
S3=V3
K10=CL/V1 K12=Q/V1 K21=Q/V2 K14=Q2/V1 K41=Q2/V3
; SC parameters F3 = THETA(5)
KA = THETA(6)
$DES
DADT(1) = K21*A(2) - K12*A(1) - K10*A(1) + KA*A(3) - K14*A(1) + K41*A(4)
DADT(2) = K12*A(1) - K21*A(2)
DADT(3) = -KA*A(3)
DADT(4) = K14*A(1) - K41*A(4)
$ERROR
IPRED=F
Y=IPRED+IPRED* ERR(1)
IRES = DV – IPRED
IWRES = IRES/IPRED
~~~

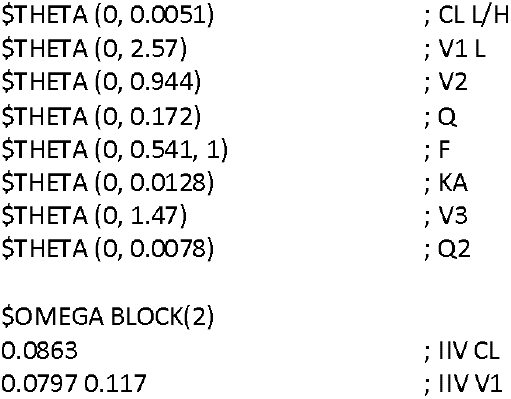

~~~
$SIGMA 0.0337
$ESTIMATION METHOD=1 INTER MAXEVAL=9999 NSIG=3 SIGL=9 PRINT=3
$COVARIANCE PRINT=E MATRIX=S
$TABLE ID GROUP TIME CMT AMT RATE DV EVID WT SEX HT AGE BLQ CL V1 V2 Q IPRED PRED CWRES CWRESI IWRES ETA1 ETA2 NOPRINT ONEHEADER NOAPPEND FILE=sdtab1
~~~

**NONMEM control stream for the final pharmacodynamic model**

~~~
$PROB TB31F
$INPUT ID GROUP SUB MOSN OOCT DV MOSNT TIME CONC EVID CMT BLQ AMT RATE SEX HT AGE WT IIN LST
$DATA TB31F_NM_PD.csv IGNORE=@
$ABBREV PROTECT
$PRED LCONC = -2
IF(CONC.NE.0) LCONC = LOG10(CONC)
EMAX = 1
EC50 = THETA(1)
HILL = THETA(2) TVBASE = THETA(3)
BASE = TVBASE * EXP(ETA(1))
DE = EMAX*(CONC**HILL)/((CONC**HILL)+(EC50**HILL))
LAMB = BASE * (1- DE)
OVDP = THETA(4) ;overdispersion factor
IF(DV.LE.1) THEN LFAC=0
ELSE
LFAC = DV*LOG(DV)-DV +LOG(DV*(1+4*DV*(1+2*DV)))/6 +LOG(3.1415)/2 ENDIF
;gamma functions of the negative binomial model expression
LGAM1=LOG(SQRT(2*3.1415))+((DV+1/OVDP)-0.5)*LOG((DV+1/OVDP))-(DV+1/OVDP)+LOG(1+1/(12*(DV+1/OVDP)))
LGAM2=LOG(SQRT(2*3.1415))+((1/OVDP)-0.5)*LOG((1/OVDP))-(1/OVDP)+LOG(1+1/(12*(1/OVDP)))
LTRM1=(LOG(1/(1+OVDP*LAMB)))*(1/OVDP)
LTRM2=(LOG(LAMB/(LAMB+1/OVDP)))*(DV)
;Logarithm of the Negative Binomial distribution
LNB = LGAM1-LFAC-LGAM2+LTRM1+LTRM2 ;Ln(negative binomial)
;-2 Log Likelihood
Y=-2*LNB
~~~

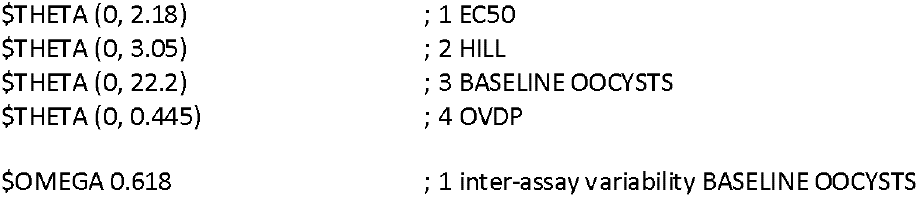

~~~
$ESTIMATION METHOD=COND LAPLACE -2LL MAXEVAL=9999 NSIG=3 SIGL=9 PRINT=3
$COVARIANCE PRINT=E MATRIX=S
$TABLE ID GROUP SUB IIN MOSN TIME CONC DV WT SEX HT AGE LAMB PRED LST ETA1 BASE LCONC NOPRINT ONEHEADER NOAPPEND FILE=sdtab1
~~~

